# Major depressive disorder, neuroticism, suicidal behaviors, and depression severity are all associated with neurotoxic immune networks and their intricate interactions with metabolic syndrome

**DOI:** 10.1101/2024.01.20.24301553

**Authors:** Michael Maes, Ketsupar Jirakran, Asara Vasupanrajit, Bo Zhou, Chavit Tunvirachaisakul, Abbas F. Almulla

## Abstract

**Background:** Major depressive disorder (MDD) is considered to be a neuroimmune disorder. However, there are no data regarding the association between comprehensive immune profiles and their interactions with the metabolic syndrome (MetS) in predicting neuroticism, suicidal behaviors, and severity of outpatient MDD (OMDD).

**Methods:** We assayed 48 serum cytokines, chemokines, and growth factors using a multiplex assay in 67 healthy controls and 66 OMDD patients. Around 50% of the OMDD and control participants had a diagnosis of MetS.

**Results:** Ten differentially expressed proteins (DEPs) were upregulated in OMDD (i.e., CXCL12, TNFB, PDGF, CCL11, IL9, IL4, CCL5, CCL2, CCL4, IL1RN), indicating an immune, defense and stress response. Six DEPs were downregulated (VEGFA, IL12, CCL3, CSF1, IL1B, NGF), indicating lowered neurogenesis and regulation of neuron death. Significant interactions between OMDD and MetS caused a) substantial increases in TNF signaling, and upregulation of IL4, IL17, TNF, TNFB, CCL2, CCL5, PDGF, IL1RN; and b) downregulation of VEGFA and FGF. A large part of the variance in neuroticism (26.6%), suicidal behaviors (23.6%), and the MDD phenome (31.4%) was predicted by immunological data and interactions between MetS and CCL5, TNFB or VEGFA.

**Discussion:** OMDD is characterized by an immunoneurotoxic profile which partly explains neuroticism, suicidal behaviors, and the phenome’s severity. Lowered IL-10 and increased neurotoxicity are characteristics of OMDD and other depression phenotypes, including severe first-episode inpatient MDD. The presence of MetS in OMDD considerably exacerbates immunoneurotoxicity. Consequently, immune studies in MDD should always be performed in subjects with and without MetS.

## Introduction

Major depressive disorder (MDD) is associated with immune system abnormalities, according to the findings of several studies (Maes, Bosmans et al. 1990, Maes, Meltzer et al. 1995, Berk, Williams et al. 2013, Maes and Carvalho 2018). Variations in immune homeostasis between the compensatory immunoregulatory system (CIRS) and the immune-inflammatory response system (IRS) are distinct manifestations of MDD (Maes, Meltzer et al. 1995, Maes, Berk et al. 2012, Maes and Carvalho 2018, Debnath, Berk et al. 2021, Maes, Rachayon et al. 2022). The IRS is the component of the immune system that initiates acute IRS activation, which is directed at addressing acute injuries, or a chronic IRS response, which develops when the acute IRS response fails to resolve (Maes, Berk et al. 2012, Maes and Carvalho 2018). The IRS is comprised of various immune profiles, including Th-17 cells, M1 macrophages, and T helper (Th)-1, in addition to the inflammatory response, which is distinguished by the acute phase response and increased production of the pro-inflammatory cytokines interleukin (IL)-1, IL-6, and tumor necrosis factor (TNF-α) (Maes, Berk et al. 2012, Maes and Carvalho 2018). The CIRS is an immune system component that functions in opposition to the IRS, thus inhibiting hyperinflammation. Additionally, it is involved in the repair phase and inflammation resolution that ensues after an acute inflammatory response (Maes, Berk et al. 2012, Maes and Carvalho 2018). This segment of the immune system is composed of negative immunoregulatory cytokines that are associated with Th-2 and T regulatory (Treg) cell lines. These cytokines include IL-4 and IL-10, which hinder the polarization of M1 and Th1, as well as certain cytokine receptors, namely soluble IL-1 receptor antagonist (sIL-1RA) and sIL-2R (Maes, Bosmans et al. 1990, Maes, Bosmans et al. 1997).

In MDD, distinct changes in IRS and CIRS components and the IRS/CIRS ratio may be observed in various phenotypes and phases of MDD (Maes and Carvalho 2018). The acute phase of severe inpatient MDD (IMDD) (including melancholia and psychotic depression) is distinguished by an increased IRS/CIRS ratio, an inflammatory response, IRS, M1 macrophage and Th-1 activation (Maes 1993, Maes and Carvalho 2018). Enhanced TNF and IL-16 signaling, in addition to T cell activation, are hallmarks of severe first episode (FE)-IMDD (Almulla, Ali Abbas Abo et al. 2023a). Furthermore, alterations in these immune profiles, cytokines, chemokines, and growth factors are indicative of the severity of MDD and its defining features, which include suicidal ideation (Maes, Rachayon et al. 2022, Almulla, Ali Abbas Abo et al. 2023a). On the contrary, very mild first episode outpatient MDD (FE-OMDD) is distinguished by a suppression of the CIRS, which results in an elevated IRS/CIRS ratio (Michael, Asara et al. 2023). Conversely, during the phase of partial remission, T regulatory activities and, consequently, CIRS activities may predominate, indicating the involvement of repair and healing processes (Maes and Carvalho 2018). However, no data exist regarding the potential association between immune profiles and key features, including neuroticism, suicidal behaviors, severity of cognitive symptoms, and the phenome, among a representative sample of OMDD patients.

In addition to distinct MDD phenotypes, metabolic syndrome (MetS) may also have an effect on immune profiles. MetS is identified through the clinical manifestations of hypertension, visceral adiposity, dyslipidemia, reduced high-density lipoprotein cholesterol (HDL-C) levels, and insulin resistance (Alberti, Eckel et al. 2009). With a prevalence of 24% in the United States population and reaching 43% among older adults, MetS increases the risk of mortality, cardiovascular disease (CVD), and type 2 diabetes mellitus (T2DM) (Wilson, Kannel et al. 1999). Furthermore, MDD and MetS exhibit a significant comorbidity (de Melo, Nunes et al. 2017), and the simultaneous presence of MDD and MetS is associated with heightened atherogenicity and insulin resistance (Nunes, Piccoli de Melo et al. 2015, de Melo, Nunes et al. 2017, Morelli, Maes et al. 2021). This is significant because increased levels of acute phase proteins, such as C-reactive protein (CRP), fibrinogen, IL-6, TNF-α, IL-18, and chemokines, and increased macrophage activation suggest that MetS is associated with a mild chronic inflammatory response (Di Lorenzo, Dell’agli et al. 2013, Zafar, Khaliq et al. 2018, Chan, Cathomas et al. 2019, Reddy, Lent-Schochet et al. 2019). While the evidence is inconclusive, some authors even classify MetS as an inflammatory disorder (Chan, Cathomas et al. 2019). The authors of this review assert that obesity is the primary cause of metabolic inflammation (Chan, Cathomas et al. 2019). However, no publications have compared the comprehensive immune networks of OMDD in terms of their key phenome features and the presence of MetS.

Therefore, the current investigation was carried out in a representative OMDD sample in order to identify alterations in the immune network, as measured by a 48-multiplex method (Maes, Rachayon et al. 2022, Almulla, Ali Abbas Abo et al. 2023a), and its associations with key features, such as suicidal ideation, neuroticism, cognitive symptoms of depression, and the depression phenome (Vasupanrajit, Maes et al. 2023). This analysis also accounted for the influence of MetS. The a priori hypotheses posit that the activated IRS profiles of OMDD and MetS are comparable, and that the interaction between OMDD and MetS induces more pronounced immune responses.

## Methods and Participants

### Participants

We included 67 healthy controls and 66 outpatients with MDD in this study. We incorporated male and female Thai-speaking participants ranging in age from 18 to 65 years. As outpatients of the Department of Psychiatry at King Chulalongkorn Memorial Hospital in Bangkok, Thailand, participants were enrolled. The control group was enlisted through oral communication within the catchment area, specifically Bangkok, between September 2021 and February 2022. Healthy volunteers included staff members, family members, associates of staff, and acquaintances of MDD patients. MDD was identified in the patients in accordance with the DSM-5 criteria (American Psychiatric Association and Association 2013). With a broad recurrence of illness (ROI) index of 1 to 20 episodes, a broad Hamilton Depression Rating Scale score range of 7 to 33 (including non-responders and partial remitters to treatment), a female to male sex ratio of 2.66, and a mean age of 36.9 (SD=11.5) years, our OMDD sample is representative of the Bangkok OMDD population.

The exclusion criteria for both patients and controls are as follows: a) psychiatric disorders falling under axis-1, which includes substance use disorders (SUD) (excluding tobacco use disorder), bipolar disorder, schizophrenia, schizo-affective psychosis, autism spectrum disorders, and psycho-organic syndromes; b) axis-2 disorders, which includes antisocial and borderline personality disorder; c) neurological disorders, which include multiple sclerosis, epilepsy, brain tumors, stroke, Alzheimer’s and Parkinson’s disorder; d) pregnant and lactating women; e) medical illnesses, which include (auto)immune disorders, rheumatoid arthritis, psoriasis, systemic lupus erythematosus, inflammatory bowel disease; f) infection one month prior to the study; g) recent surgery; h) subjects who were ever treated with immunomodulatory drugs; i) subjects who took therapeutically doses of antioxidants and omega-3 three months prior to the investigation; and j) frequent use of pain killers. Furthermore, controls who presented with MDD (both current and lifetime) and DSM-IV-TR anxiety disorders, as well as a positive familial history of mood disorders, suicide, and substance use disorders, were precluded from the study.

Before participating in the study, written informed consent was voluntarily obtained from all subjects. The research was carried out in adherence to privacy laws and international and Thai ethical standards. The investigation was authorized (#445/63) by the Institutional Review Board of Chulalongkorn University’s Faculty of Medicine in Bangkok, Thailand.

### Clinical measurements

Sociodemographic and clinical information were collected via interviews with patients and controls using a semi-structured questionnaire. Sex, marital status, employment status, occupation, income, year of education, family history of substance use disorders, marital status, and employment status, are all components of the semi-structured interview. Additionally, the interview inquires about the use of psychotropic medications and medical history. The DSM-5 criteria and the Mini International Neuropsychiatric Interview (M.I.N.I.) were utilized to diagnose MDD (Kittirattanapaiboon 2004). The Beck Depression Inventory II (BDI-II) (Beck, Steer et al. 1996) and the Hamilton Rating Scale for Depression (HAM-D) (Hamilton 1960) were utilized to assess the severity of MDD. The BDI-II is a self-report inventory consisting of 21 items. A Thai translation of the inventory was used (Mungpanich 2008). A pure cognitive depression BDI score was calculated by aggregating the 16 cognitive depressive items of the BDI. In doing so, the items measuring appetite, weight loss, fatigue, irritability, insomnia, and sex drive were omitted. The Columbia Suicide Severity Rating Scale (C-SSRS) (Posner, Brown et al. 2011) was employed to evaluate the severity of past and present suicidal ideation and attempts. The severity, intensity, lethality, and frequency of suicidal ideation (SI) and attempts (SA) were assessed using the C-SSRS. A Thai translation of the Big Five Inventory (BFI) (John and Srivastava 1999) was utilized to evaluate five personality traits (Luangsurong 2016). The raw scores of the neuroticism dimension were utilized in the current investigation, given its significant contribution to the MDD phenome (Jirakran, Vasupanrajit et al. 2023).

Indices were developed in accordance with the rating scale assessments to represent the OMDD phenome. To calculate the Recurrence of Illness (ROI) index, the first principal component (PC) was derived from the number of depressive episodes, past suicidal attempts, and suicidal ideation. Maes et al. (Michael, Ketsupar et al. 2023) calculated the current suicidal behavior score by utilizing the first PC extracted from seven items measuring current suicidal ideation on the C-SSRS and the first PC extracted from five items measuring suicidal attempts on the C-SSRS. Subsequently, current suicidal behavior was represented by the aggregate of the z-transformations of these two PC scores. Maes et al. (Michael, Ketsupar et al. 2023) defined the severity of the current OMDD phenome as the first PC derived from the cognitive BDI score, HAM-D, and current suicidal behaviors. Each of these PCs met rigorous predetermined standards (refer to the Statistics).

The formula utilized to determine body mass index (BMI) was weight (kg) divided by height (m) squared. MetS was defined as the presence of three or more of the following components in accordance with the 2009 Joint Scientific Statement of the American Heart Association/National Heart, Lung, and Blood Institute (Alberti, Eckel et al. 2009): (a) A waist circumference of 90 cm or greater for men and 80 cm for women, or a body mass index (BMI) of >25 kg/m2; (b) A high triglyceride level of 150 mg/dL; (c) A low HDL cholesterol level of <40 mg/dL for men and <50 mg/dL for women; (d) high blood pressure of >130 mm Hg systolic blood pressure, 85 mm Hg diastolic blood pressure, or antihypertensive medication; e) increase fasting glucose (≥ 100 mg/dL) or a diabetes diagnosis. To investigate the effect of MetS on immune profiles, we comprised approximately 50% of the participants in the control and MDD samples with MetS. As a result, 64 participants had MetS and 69 did not have the condition. The distribution of MetS in MDD patients and controls is displayed in **Table 1**. The DSM-5 criteria were applied in the diagnostic process of tobacco use disorder (TUD).

**Table 1.** Demographic and clinical data of the patients with major depression (MDD) and healthy controls (HC) in the current study.

| Variables | HC (n=67) | MDD (n=66) | F/X2 | df | P |
| --- | --- | --- | --- | --- | --- |
| Age (years) | 37.9 (9.2) | 37.0 (11.5) | 0.26 | 1/131 | 0.609 |
| Sex (F/M) | 58/9 | 48/18 | 3.94 | 1 | 0.055 |
| Education (years) | 14.1 (3.4) | 14.8 (2.8) | 1.72 | 1/131 | 0.193 |
| Number of episodes | - | 3.9 (3.9, range: 1-20) | - | - | - |
| SBP | 127.9 (15.3) | 129.9 (17.8) | 0.48 | 1/131 | 0.492 |
| DBP | 79.2 (11.3) | 77.9 (12.4) | 0.42 | 1/131 | 0.518 |
| BMI (kg/m2) | 27.4 (6.1) | 26.9 (5.4) | 0.27 | 1/130 | 0.607 |
| MetS (No/Yes) | 34/33 | 35/31 | 0.07 | 1 | 0.792 |
| Smoking (No/Yes) | 62/5 | 54/12 | 3.43 | 1 | 0.064 |
| Total HAM-D score | 2.3 (3.0) | 18.1 (6.4) | 330.94 | 1/130 | <0.001 |
| Total BDI-II score | 5.6 (6.5) | 26.9 (13.8) | 129.52 | 1/130 | <0.001 |
| Pure BDI (z score) | -0.687 (0.376) | 0.697 (0.952) | 122.20 | 1/130 | <0.001 |
| ROI (z score) | -0.752 (0.0) | 0.776 (0.918) | MWU | - | <0.001 |
| Neuroticism (z score) | -0.609 (0.686) | 0.619 (0.883) | 80.33 | 1/130 | <0.001 |
| SB (z score) | -0.554 (0.0) | 0.571 (1.181) | 60.74 | 1/130 | <0.001 |
| MDD Phenome (z score) | -0.749 (0.421) | 0.760 (0.828) | 175.97 | 1/130 | <0.001 |

All data are shown as mean (SD) or as ratio. All results of analysis of variance (F values) or analysis of contingency analysis ( $X^2$ test). MWU: Mann-Whitney U test.
SBP and DBP: systolic and diastolic blood pressure, BMI: body mass index, MetS: metabolic syndrome, HAM-D: Hamilton Depression Rating Scale score, BDI: Bech Depression Inventory, ROI: recurrence of illness index, SB: suicidal behaviors.

### Assays

Using a serum tube and disposable syringe, we collected 5 milliliters of fasting venous blood from each participant in the study between 8:00 and 9:00 a.m. After centrifugation of blood at 35,000 rpm, the serum was collected and preserved in Eppendorf tubes with aliquots at −80 °C, until it was thawed for the purpose of conducting biomarker assays. The fluorescence intensities (FI) of forty-eight cytokines, chemokines, and growth factors were measured using the Bio-Plex Multiplex Immunoassay kit from Bio-Rad Laboratories Inc., Hercules, USA. Table 1 of the Electronic Supplementary File (ESF) contains a comprehensive inventory of the analytes utilized in our research, including their corresponding gene IDs and aliases. Our methodology comprised the subsequent stages: a) Using sample diluent (HB), the serum was diluted to 1:4. b) The samples, once diluted, were transferred to a 96-well plate, each well containing 50µl of a microparticle cocktail (comprising cytokines, chemokines, and growth factors). Subsequently, the plate underwent incubation at room temperature with shaking at 850 rpm for one hour. C) After three washes, each well of the plate received 50µl of diluted Streptavidin-PE, followed by a ten-minute shaking at room temperature. d) After the sample transfer to the 96-well plate, the subsequent analysis for evaluating cytokines, chemokines, and growth factors was performed using the Bio-Plex® 200 System from Bio-Rad Laboratories, Inc. The intra-assay CV values for every analyte were found to be below 11.0%. The concentrations of the samples were ascertained using the standard concentrations provided by the manufacturer. The percentage of concentrations below the lowest measurable concentration (out of range or OOR) was subsequently assessed. The number of analytes that exhibited measurable values (i.e., those surpassing OOR) is detailed in ESF, Table 1. We did not enter analytes with more than 60% OORs in the analyses, and consequently, IL-2, IL-3, IL-5, IL-6, IL-7, and IL-12p70 were omitted. Analytes that showed quantifiable levels spanning from 40% to 60% were regarded as prevalences. Consequently, IL-10 was substituted as a dummy variable. All other variables were used as scale variables. Blank-subtracted FI values were utilized in our statistical analysis as opposed to absolute concentrations because they are more appropriate, especially when multiple plates are utilized. ESF, Table 2 shows the immune profiles that were computed as z unit-based composite scores (Maes and Carvalho 2018, Almulla, Ali Abbas Abo et al. 2023a).

**Table 2.** Results of logistic binary regression analysis with major depressive episode as dependent variable (and controls as reference group) in participants with and without metabolic syndrome (MetS).

| Samples | Explanatory variables | B | SE | Wald | df | p | OR | Lower 95% | Upper 95% |
| --- | --- | --- | --- | --- | --- | --- | --- | --- | --- |
| #1. ALL | HGF | -0.745 | 0.262 | 8.09 | 1 | 0.004 | 0.475 | 0.284 | 0.793 |
|  | CXCL12 | 1.248 | 0.280 | 19.89 | 1 | <0.001 | 3.483 | 2.013 | 6.028 |
| | TNF-β | 1.103 | 0.277 | 15.85 | 1 | <0.001 | 3.013 | 1.750 | 5.187 |
|  | VEGF | -0.833 | 0.253 | 10.82 | 1 | 0.001 | 0.435 | 0.265 | 0.714 |
| #2. No MetS | CCL11 | 1.023 | 0.340 | 9.05 | 1 | 0.003 | 2.780 | 1.428 | 5.414 |
|  | LIF | 0.774 | 0.339 | 5.22 | 1 | 0.022 | 2.168 | 1.116 | 4.209 |
|  | M-CSF | -1.485 | 0.435 | 11.68 | 1 | 0.001 | 0.227 | 0.097 | 0.531 |
| #3. MetS | CCL5 | 3.413 | 1.112 | 9.417 | 1 | 0.002 | 30.37 | 3.262 | 268.631 |
| | TNF-β | 1.823 | 0.795 | 5.255 | 1 | 0.022 | 6.19 | 1.303 | 29.418 |
|  | VEGF | -2.006 | 0.790 | 6.447 | 1 | 0.011 | 0.135 | 0.029 | 0.633 |
| #4. ALL | PC05 x MetS | 2.096 | 0.521 | 16.173 | 1 | <0.001 | 8.132 | 2.928 | 22.582 |
|  | PCinv x MetS | -1.265 | 0.413 | 9.400 | 1 | 0.002 | 0.282 | 0.126 | 0.634 |
| #5. ALL | CCL5 x MetS | 3.4298 | 0.937 | 12.398 | 1 | <0.001 | 27.050 | 4.314 | 169.615 |
| | TNF-β x MetS | 1.743 | 0.678 | 6.616 | 1 | 0.010 | 5.717 | 1.514 | 21.584 |
|  | VEGF x MetS | -1.564 | 0.602 | 6.759 | 1 | 0.009 | 0.209 | 0.064 | 0.680 |
|  | CCL11 | 0.758 | 0.260 | 8.484 | 1 | 0.004 | 2.134 | 1.281 | 3.553 |
|  | M-CSF | -0.808 | 0.285 | 8.037 | 1 | 0.005 | 0.446 | 0.255 | 0.779 |

OR: Odds ratio, 95% CI: 95% confidence intervals. For cytokine abbreviations: see ESF, Table 1, MetS: metabolic syndrome, X: interaction terms.

### Statistical analysis

For comparing nominal variables across different categories, either the chi-square test or Fisher’s Exact Probability Test was employed, while analysis of variance was used for assessing scale variables among various diagnostic groups. The correlations between two sets of scale variables were calculated utilizing Pearson’s product moment or Spearman’s rank order coefficients. Point-biserial correlation coefficients were employed to investigate the associations between scale and binary variables. When analyzing the immune data, no false discovery rate (FDR) p-correction was applied (to multiple comparisons or correlations) (Maes, Rachayon et al. 2022). In fact, due to their tight networks of interactions, serum cytokines, chemokines, and growth factors cannot be considered independent variables in statistical analyses. PC analysis (PCA) was conducted to reduce the number of variables into a single PC score, which was subsequently applicable to additional statistical analyses. The Bartlett’s sphericity test and the Kaiser-Meyer-Olkin test for sample adequacy (which yields satisfactory results between 0.5 and 0.6), were employed to ascertain factorability. To construct reflective clinical PC models (e.g., OMDD phenome, ROI, suicidal behaviors), the first PC was deemed acceptable only if the variance explained (VE) exceeded 50%, all loadings on the initial PC exceeded 0.6, and Cronbach’s alpha exceeded 0.7. However, we also calculated the first PCs derived from immune measurements and permitted three PC loading levels: 0.4, 0.5, and 0.6. A reduced VE was acceptable when Cronbach’s alpha was greater than 0.9 and protein-protein interaction (PPI) analysis indicated that the PC’s cytokines, chemokines, and growth factors also constituted a dense network (as determined by the average clustering coefficient).

Using multiple regression analysis (manual method), the effects of explanatory variables (immune data) on dependent variables (clinical data) were investigated. Furthermore, to determine the most accurate predictors for the model, we employed a forward stepwise automatic regression technique with p-values of 0.05 to enter and 0.06 to eliminate. The final regression models incorporated standardized coefficients, t-statistics, and exact p-values for each of the explanatory variables, in addition to F statistics (and p values) and total variance (R^2^ or partial eta squared as effect size) that were accounted for by the model. Tolerance (cut-off value <0.25), the variance inflation factor (cut-off value >4), the condition index, and variance proportions from the collinearity diagnostics table were employed to analyze multicollinearity and collinearity. To ascertain the presence of heteroskedasticity, the White and modified Breusch-Pagan test was applied. With OMDD as the dependent variable and no-OMDD as the reference group, a binary logistic regression analysis was conducted; the exploratory variables were immune data and putative confounders. The computation of the odds ratio (OR), 95% confidence intervals (95% IC), accuracy, confusion matrix, and Nagelkerke pseudo-R^2^ effect size were performed. Each of the studies mentioned above utilized a two-tailed design, and statistical significance was established at an α value of 0.05. As required, the data distributions were normalized using logarithmic, square-root, or rank-based, inversed normal transformations. Additionally, the data underwent z-score transformation to enhance their interpretability and to generate z-unit-weighted composite scores that more accurately represent distinct immune profiles. Version 29 of IBM Windows SPSS was utilized. G*Power 3.1.9.4 was employed to calculate the a priori sample size. The primary statistical analysis that was predetermined was a multiple regression analysis, in which the immune data and phenome score were utilized as input variables and output variables, respectively. The minimal sample size required, considering f = 0.136 (approximately 12% explained variance), number of explanatory variables = 5, alpha = 0.05, and power = 0.8, was 100.

We assessed zero-order protein-protein interaction (PPI) networks utilizing STRING version 12.0 (https://string-db.org, accessed 12-01-2023) and GoNet (https://tools.dice-database.org/Gonet/, accessed 12-01-2023). The set organism “homo sapiens” and a minimal required interaction score of 0.400 were both specified. We outlined the network characteristics using STRING, which comprised the number of nodes and edges, expected number of edges, average node degree, and average clustering coefficient. Annotation and enrichment analysis were performed utilizing STRING and GoNet; the networks were subsequently queried against the Gene Ontology (GO) database of biological processes. STRING was utilized to generate interactive diagrams displaying the selected GO annotations and the differentially expressed protein (DEP) network.

## Results

### Sociodemographic data of patients and controls

**Table 1** shows the sociodemographic and clinical assessments in MDD outpatients and healthy controls. There were no significant differences in age, male/female ratio, education, blood pressure, BMI, prevalence of MetS, and TUD between patients and controls. The total HAM-D and BDI-II scores and the pure BDI-II scores were significantly higher in patients than controls. ROI, neuroticism scores, suicidal behavior and phenome scores were all higher in MDD patients than in controls.

### Immune profiles in MDD patients and controls

ESF, Table 3a and 3b show the measurements of the immune profiles in both MDD patients versus controls, and those with and without MetS. The data were analyzed using univariate GLM analysis with MDD and MetS as fixed factors while allowing for the interaction patterns between MDD x MetS and for the effects of age and sex (and BMI in the case we looked for the differences between MDD, but not MetS, and controls). There were no significant differences in M1, M2, Th-1, Th-2, IRS, and CIRS between MDD patients and controls and between subjects with and without MetS. IL-1 signaling was significantly lower in MDD than in controls, whereas TNF signaling was significantly increased in MDD versus controls. There were no significant changes in any of the immune profiles between those with and without MetS.

**Table 3.** Multiple regression analysis with clinical phenome data as dependent variables and cytokines/chemokine/growth factors as explanatory variables.

| Dependent Variables | Explanatory Variables | Coefficients of input variables |  |  | Model statistics |  |  |  |
| --- | --- | --- | --- | --- | --- | --- | --- | --- |
| | | $\beta$ | t | p | R<sup>2</sup> | F | df | p |
| #1. ROI | Model |  |  |  | 0.276 | 12.09 | 4/127 | <0.001 |
| | TNF-β | 0.322 | 3.84 | <0.001 | | | | |
|  | CXCL12 | 0.333 | 4.12 | <0.001 |  |  |  |  |
|  | VEFG | -0.209 | -2.68 | 0.008 |  |  |  |  |
| #2. Neuroticism | Model |  |  |  | 0.155 | 7.90 | 3/129 | <0.001 |
| | TNF-β | 0.183 | 2.18 | 0.031 | | | | |
|  | VEGF | -0.268 | -3.23 | 0.002 |  |  |  |  |
|  | CXCL12 | 0.215 | 2.53 | 0.013 |  |  |  |  |
| #3. LT SI+SA | Model |  |  |  | 0.102 | 7.31 | 2/129 | 0.001 |
|  | CCL5 | 0.307 | 3.58 | <0.001 |  |  |  |  |
|  | M-CSF | -0.179 | -2.10 | 0.038 |  |  |  |  |
| #4. Pure BDI-II | Model |  |  |  | 0.215 | 8.77 | 4/128 | <0.001 |
| | TNF-β | 0.262 | 3.26 | 0.001 | | | | |
|  | VEFG | -0.239 | -2.86 | 0.005 |  |  |  |  |
|  | IL-12p40 | -0.183 | -2.22 | 0.028 |  |  |  |  |
| | IFN-γ | 0.177 | 2.18 | 0.031 | | | | |
| #5. Phenome | Model |  |  |  | 0.265 | 7.58 | 6/126 | <0.001 |
| | TNF-β | 0.233 | 2.91 | 0.004 | | | | |
|  | VEGF | -0.273 | -3.21 | 0.002 |  |  |  |  |
| | IFN-γ | 0.186 | 2.20 | 0.030 | | | | |
|  | M-CSF | -0.239 | -2.83 | 0.005 |  |  |  |  |
|  | IL-18 | 0.218 | 2.51 | 0.013 |  |  |  |  |
|  | IL-12p40 | -0.169 | -2.10 | 0.037 |  |  |  |  |
| #6. ROI | Model |  |  |  | 0.390 | 16.14 | 5/126 | <0.001 |
|  | CCL5 x MetS | 0.276 | 3.36 | 0.001 |  |  |  |  |
| | TNF-β | 0.279 | 3.58 | <0.001 | | | | |
|  | HGF | -0.247 | -3.30 | 0.001 |  |  |  |  |
|  | CXCL12 | 0.228 | 2.84 | 0.005 |  |  |  |  |
|  | PCinv x MetS | -0.203 | -2.80 | 0.006 |  |  |  |  |
| **#7. Neuroticism** | **Model** |  |  |  | 0.266 | 9.21 | 5/127 | <0.001 |
| | TNF-β x MetS | 0.245 | 3.00 | 0.003 | | | | |
|  | PCinv | -0.333 | -3.90 | <0.001 |  |  |  |  |
|  | CXCL12 | 0.182 | 2.30 | 0.023 |  |  |  |  |
|  | LIF | 0.196 | 2.33 | 0.021 |  |  |  |  |
|  | M-CSF | -0.164 | -2.13 | 0.035 |  |  |  |  |
| **#8. Suicidal behaviors** | **Model** |  |  |  | 0.236 | 9.80 | 4/127 | <0.001 |
|  | CCL5 x MetS | 0.267 | 2.87 | 0.005 |  |  |  |  |
|  | VEGF x MetS | -0.212 | -2.60 | 0.010 |  |  |  |  |
|  | M-CSF | -0.179 | -2.27 | 0.025 |  |  |  |  |
|  | CXCL12 | 0.196 | 2.20 | 0.030 |  |  |  |  |
| **#9. Phenome** | **Model** |  |  |  | 0.314 | 9.60 | 6/126 | <0.001 |
|  | TNF signaling x MetS. | 0.211 | 2.48 | 0.014 |  |  |  |  |
|  | PCinv | -0.268 | -3.38 | 0.001 |  |  |  |  |
|  | CCL5 x MetS | 0.179 | 2.14 | 0.035 |  |  |  |  |
|  | M-CSF | -0.259 | -3.21 | 0.002 |  |  |  |  |
| | IFN-γ | 0.184 | 2.22 | 0.028 | | | | |

Since the classical immune profiles did not differ between controls and MDD patients we have examined the immune profiles that can be retrieved in the data set. Towards that end we performed PCA to detect patterns in the immune data set. ESF, Table 4 shows a first PC (PC04) extracted from the cytokines that showed loadings > 0.4 on the first factor. The KMO index was sufficient, Cronbach’s alpha=0.914, and this first PC explained 34.17% of the variance in the data set. This PC1 loaded “highly” (>0.4) on 27 cytokines (see ESF, Table 4a) and the 27 cytokines shaped a tight PPI network (average clustering coefficient = 0.896). We constructed a second PC (PC05) that we extracted from the cytokine data with the criterion that the PC loadings should be higher than 0.5. This PC1 loaded “highly” (>0.5) on 20 cytokines, Cronbach’s alpha was adequate (see ESF, Table 4a). The KMO was sufficient, and this PC explained 40.64% of the variance in the data set while Cronbach’s alpha was 0.914 and the PPI clustering coefficient was 0.913. A third PC was constructed (PC06) based on PC loadings higher than 0.6 (ESF, Table 4c). The KMO was sufficient, Cronbach’s alpha = 0.890, while the variance explained was 54.42%. Moreover, the PPI clustering coefficient was 0.927. As such, PC06 may be regarded as a validated PC, whereby the DEPs are reflective manifestations of the PC06 construct. Nevertheless, PC04 and PC05 show that the cytokines belonging to these PCs are interrelated and are upregulated in concert. We constructed also a fourth PC (PCinv) based on the remaining cytokines and found that this PC showed Cronbach’s alpha of 0.716 and explained 33.19% of the variance. The clustering coefficient was 0.9. ESF, Tables 5a and 5b show the results of a multivariate GLM that examines the associations between MDD and the four PC scores, while adjusting for MetS, age and sex. We found that PC04, PC05, and PC06 were significantly higher in MDD than in controls, whereas PCinv was significantly lower in patients than controls. There were no significant differences in the PC scores between those with and without MetS (see ESF, Table 5c).

**Table 4.**
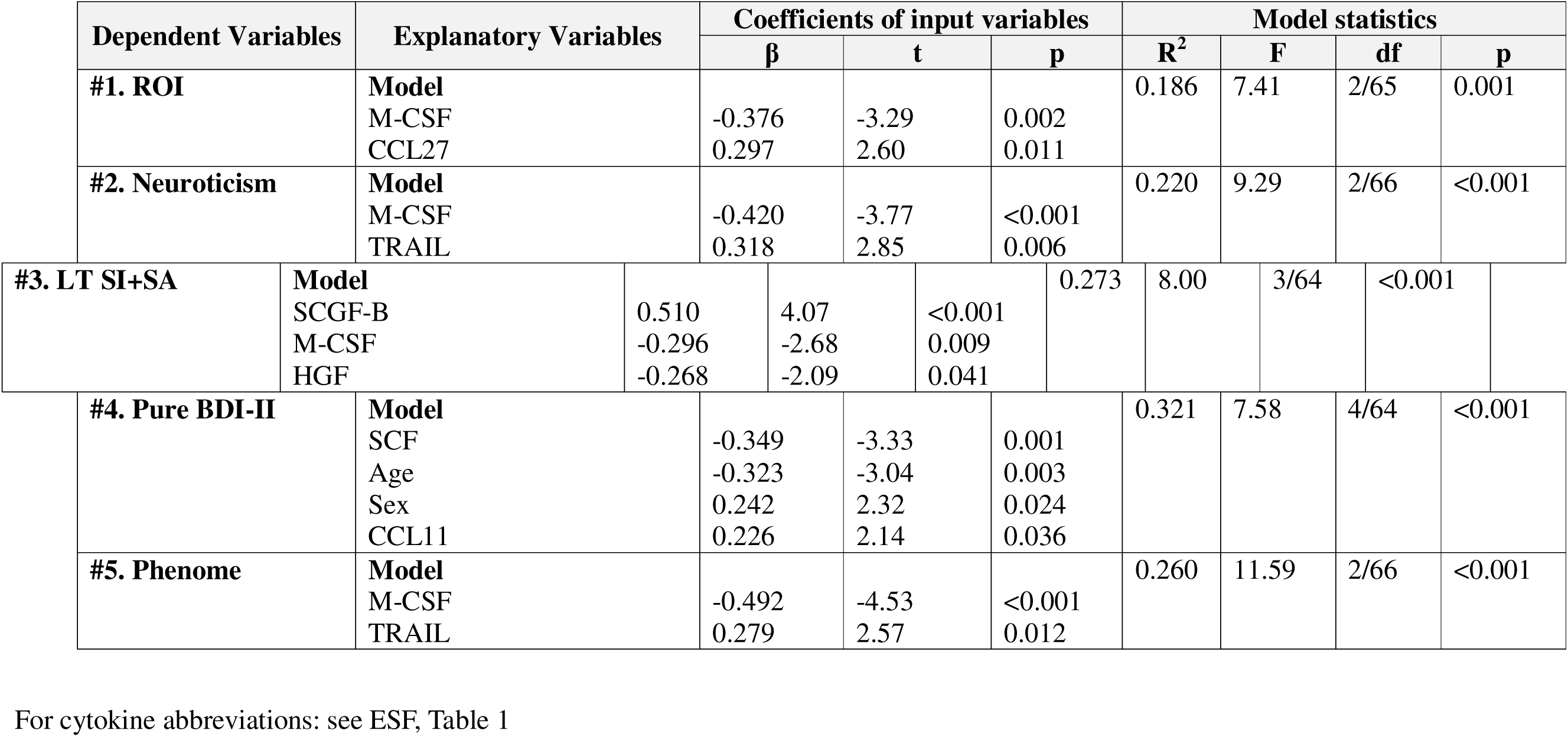
Multiple regression analysis with clinical phenome data as dependent variables and cytokines/chemokine/growth factors as explanatory variables in individual without metabolic syndrome.

**Table 5.** Multiple regression analysis with clinical phenome data as dependent variables and cytokines/chemokine/growth factors as explanatory variables in individual with metabolic syndrome.

| Dependent Variables | Explanatory Variables | Coefficients of input variables |  |  | Model statistics |  |  |  |
| --- | --- | --- | --- | --- | --- | --- | --- | --- |
| | | $\beta$ | t | p | R<sup>2</sup> | F | df | p |
| #1. ROI | **Model** |  |  |  | 0.656 | 38.51 | 3/60 | <0.001 |
|  | CCL5 | 0.322 | 3.84 | <0.001 |  |  |  |  |
|  | VEGF | 0.333 | 4.12 | <0.001 |  |  |  |  |
| | TNF-β | -0.209 | -2.67 | <0.001 | | | | |
| #2. Neuroticism | **Model** |  |  |  | 0.433 | 15.27 | 3/60 | <0.001 |
|  | sIL-1RA | 0.418 | 4.17 | <0.001 |  |  |  |  |
|  | VEGF | -0.319 | -3.16 | 0.002 |  |  |  |  |
| | TNF-β | 0.235 | 2.26 | 0.028 | | | | |
| #3. LT SI+SA | **Model** |  |  |  | 0.320 | 9.41 | 3/60 | <0.001 |
|  | CCL5 | 0.290 | 2.48 | 0.016 |  |  |  |  |
|  | VEGF | -0.417 | -3.44 | 0.001 |  |  |  |  |
|  | MIF | 0.267 | 2.11 | 0.039 |  |  |  |  |
| #4. Pure BDI-II | **Model** |  |  |  | 0.519 | 21.55 | 3/60 | <0.001 |
|  | sIL-1RA | 0.431 | 4.65 | <0.001 |  |  |  |  |
|  | VEGF | -0.376 | -4.02 | <0.001 |  |  |  |  |
| | TNF-β | 0.250 | 2.60 | 0.012 | | | | |
| #5. Phenome | **Model** |  |  |  | 0.600 | 22.10 | 4/59 | <0.001 |
|  | sIL-1RA | 0.291 | 2.64 | 0.011 |  |  |  |  |
|  | VEGF | -0.488 | -4.99 | <0.001 |  |  |  |  |
| | IFN-γ | 0.296 | 2.41 | 0.019 | | | | |
| | TNF-β | 0.194 | 2.06 | 0.044 | | | | |

For cytokine abbreviations: see ESF, Table 1

ESF, Table 6a shows the measurements of the cytokines in MDD versus controls. **Figure 1** shows a bar graph with the cytokines that were significantly increased (in descending order of importance CXCL12, TNF-β, PDGF, CCL11, IL-9, IL-4, TNF signaling, CCL2, CCL4 and sIL-1RA. The same figure also shows the cytokines that were decreased in MDD versus controls (in ascending order of importance: IL-1 signaling, NGF, IL-1β, M-CSF, CCL3, IL-12p40, VEGF. The same Figure also shows PC05 (increased) and PCinv (decreased in MDD). It should be added that the prevalence of measurable IL-10 levels was significantly lower (X^2^=6.16, df=1, p=0.013) in MDD (45.3%) than in controls (66.6%).

**Figure 1.**
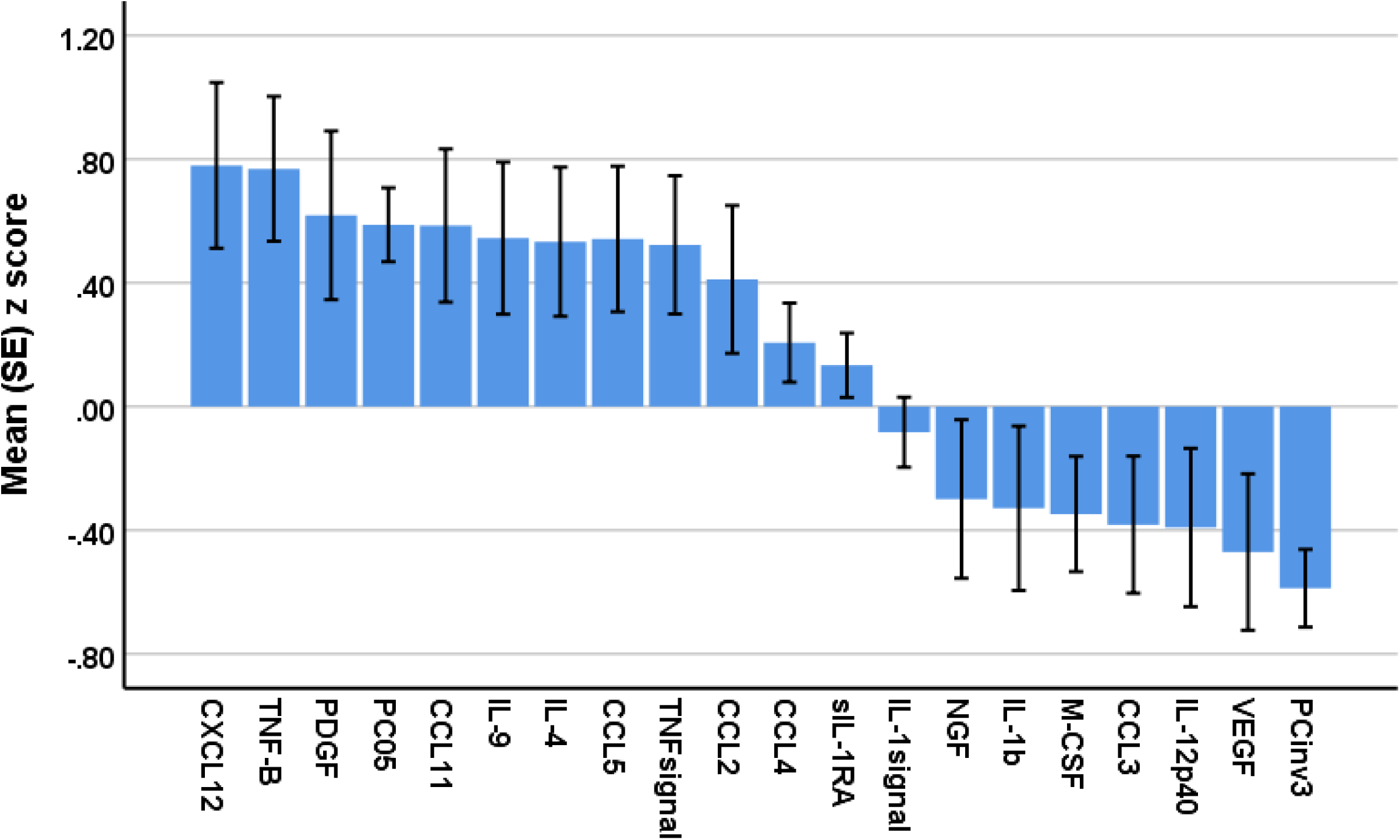
Measurements of the cytokines, chemokins, and growth factors that are upregulated and downregulated in major depression. See ESF, Table 1 for explanation of the immune data. PC05 and PCinv: principal components, see ESF, Table 4 for explanations.

**Table 6.** Intercorrelation matrix between the features of major depression and immune factors and well as their interactions with metabolic syndrome (MetS).

| **Variables** | **ROI** | **Neuroticism** | **SB** | **Pure BDI** | **Phenome** |
| --- | --- | --- | --- | --- | --- |
| **PC04** | 0.170 | 0.123 | 0.161 | 0.064 | 0.130 |
| **PC04 x MetS** | 0.289** | 0.218* | 0.254** | 0.211* | 0.257** |
| **PC05** | 0.285** | 0.239** | 0.239** | 0.184 | 0.244** |
| **PC05 x MetS** | 0.416** | 0.337** | 0.365** | 0.336** | 0.366** |
| **PCinv** | -0.245** | -0.308** | -0.163 | -0.315** | -0.293** |
| **PCinv x MetS** | -0.294** | -0.261** | -0.234** | -0.298** | -0.266** |
| **MetS** | -0.058 | -0.068 | -0.033 | -0.028 | -0.043 |
| **BMI** | -0.106 | -0.044 | -0.027 | -0.001 | 0.021 |

PC: principal component, MetS: metabolic syndrome, BMI: body mass index.
\* $P < 0.05$ , \*\* $p < 0.001$ . All results of Pearson's product moment correlation coefficients, except for MetS: results of point-biserial correlation coefficient.

A subset of the patients was prescribed psychotropic medications, such as mirtazapine (n=8), benzodiazepines (n=36), sertraline (n=28), trazodone (n=16), fluoxetine (n=7), venlafaxine (n=13), paroxetine (n=2), agomelatine (n=2), venlafaxine (n=13), and agomelatine (n=2). However, these drugs did not exhibit any discernible impact on the immune markers, either the immune PC profiles or the solitary cytokines, chemokine, or growth factors (even without p-correction).

ESF, Table 6b shows the measurements of all cytokines in those with and without MetS. No significant differences could be found between both groups in any of the cytokines. ESF, Table 6c (results of univariate GLM with MDD and MetS as fixed factors and age and sex as covariates) shows the associations between the cytokines and BMI. We found associations between BMI and 13 cytokines. Nevertheless, most associations showed a very low effect size, except sIL-1RA (R^2^=0.198), HGF (R^2^=0.0865) and TRAIL (R^2^=0.084). The other cytokines showed effect sizes < 0.06.

### Prediction of MDD using cytokines

**Table 2** shows the outcome of binary regression analyses with the diagnosis MDD as dependent variable (and controls as reference group) and immune data as explanatory variables while allowing for the effects of age, sex, education, smoking, and BMI (these were all non-significant). The analyses were re-run in three different conditions, namely all subjects combined, and in participants (controls and patients) with and without MetS. In all participants (thus with and without MetS) we found that MDD was best predicted by 4 immune variables, namely CXCL12 and TNF-β (both positively), HGF and VEGF (both inversely) (Χ^2^=59.660, df=4, p<0.001, Nagelkerke=0.482, accuracy=75.2%, sensitivity=68.2% and specificity=82.1%). None of the putative confounding factors was significant. Regression #2 shows the outcome of a binary regression analysis performed in participants without MetS. The results were quite different from those in the total group (regression #1) and showed that CCL11 and LIF increased the Odds of MDD, whereas M-CSF decreased the Odds (Χ^2^=28.97, df=3, p<0.001, Nagelkerke=0.457, accuracy=79.7%, sensitivity=82.9% and specificity=76.5%). In people with the MetS, we detected that CCL5 and TNF-β were both positively associated with MDD, whereas VEGF was inversely associated with MDD (Χ^2^=64.014, df=3, p<0.001, Nagelkerke=0.842, accuracy=92.2%, sensitivity=87.1% and specificity=97.0%).

### Prediction of the key features of MDD using immune markers

**Table 3** examines the associations between the features of MDD and the immune variables in all participants combined (with and without MetS). Again, we allow for the effects of age, sex, smoking, and BMI. Regressions #1 and #2 show that 27.6% and 15.5% of the variance in the ROI index and neuroticism was explained by CXCL12 and TNF-β (both positively associated) and VEGF (inversely associated). Suicidal behaviors (regression #3) were associated with CCL5 (positively) and M-CSF (inversely). Up to 21.5% of the variance in pure depressive symptoms (BDI-II) was explained by 4 immune variables, namely TNF-β and IFN-γ (positively) and VEGF and IL-12p40 (both inversely). The phenome score was best predicted (26.5% of the variance) by TNF-β, IFN-γ and IL-18 (all positively) and VEGF, M-CSF and IL-12p40 (both inversely).

**Table 4** shows the results of multiple regression analyses of the same dependent variables on the immune data but conducted in participants without MetS. The most significant explanatory variables are quite different from those obtained in the total study group, although the effect sizes are quite comparable. For example, in subjects without MetS, ROI was associated with lowered M-CSF but increased CCL27. We found that 22.0% of the variance in neuroticism was explained by M-CFS (decreased) and TRAIL (increased) (regression #3). Up to 27.3% of the variance in suicidal behaviors was explained by SCGF-B (positively), HGF and M-CSF (both inversely). Regression #4 shows that 32.1% of the variance in self-rated severity of depressive symptoms was explained by CCL11 (positively), SCF and age (both inversely) and male sex. The two best predictors of the phenome of MDD were TRAIL and M-CSF which together explained 26.0% of its variance.

**Table 5** shows the results of multiple regression analyses of the same dependent variables on the immune data but conducted in participants with MetS. The first striking difference with the results obtained in people without MetS is that the effects sizes are much higher. For example, 65.5% of the variance in ROI in participants with MetS could be explained by CCL5, TNF-β and VEGF, whereas in those without MetS only 18.6% of the variance was explained by CCL27 and M-CSF. **Figure 2** shows the partial regression of ROI on serum CCL5 levels. Likewise, 43.3% of the variance in neuroticism (regression #2) was explained by sIL-1RA and TNF-β (both positively) and VEGF (inversely). There was a strong association (regression #4) between suicidal behaviors and CCL5 and MIF (positively) and VEGF (inversely). A very large part of the variance in the cognitive symptoms of MDD (regression #4) was predicted by sIL-1RA, TNF-β (positively) and VEGF (inversely). Regression #5 shows that 60.0% of the variance in the phenome’s severity could be explained by the regression on sIL-1RA, IFN-γ, TNF-β (positively), and VEGF (inversely). **Figure 3** shows the partial regression of the phenome of OMDD on the PC05 score.

**Figure 2.**
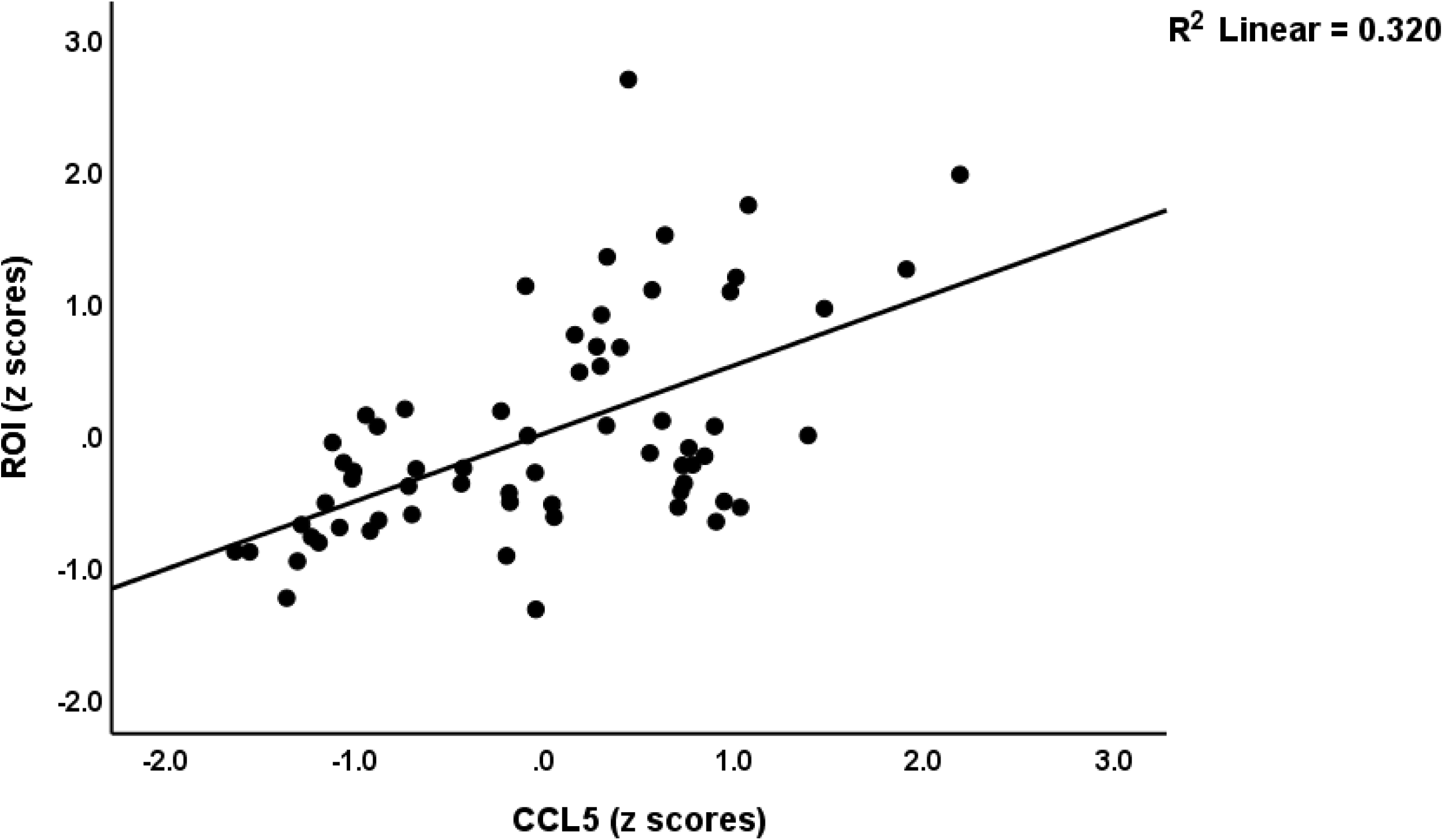
Partial regression of the recurrence of illness index (ROI) of major depression on serum CCL5 le participants with metabolic syndrome (p<0.001).

**Figure 3.**
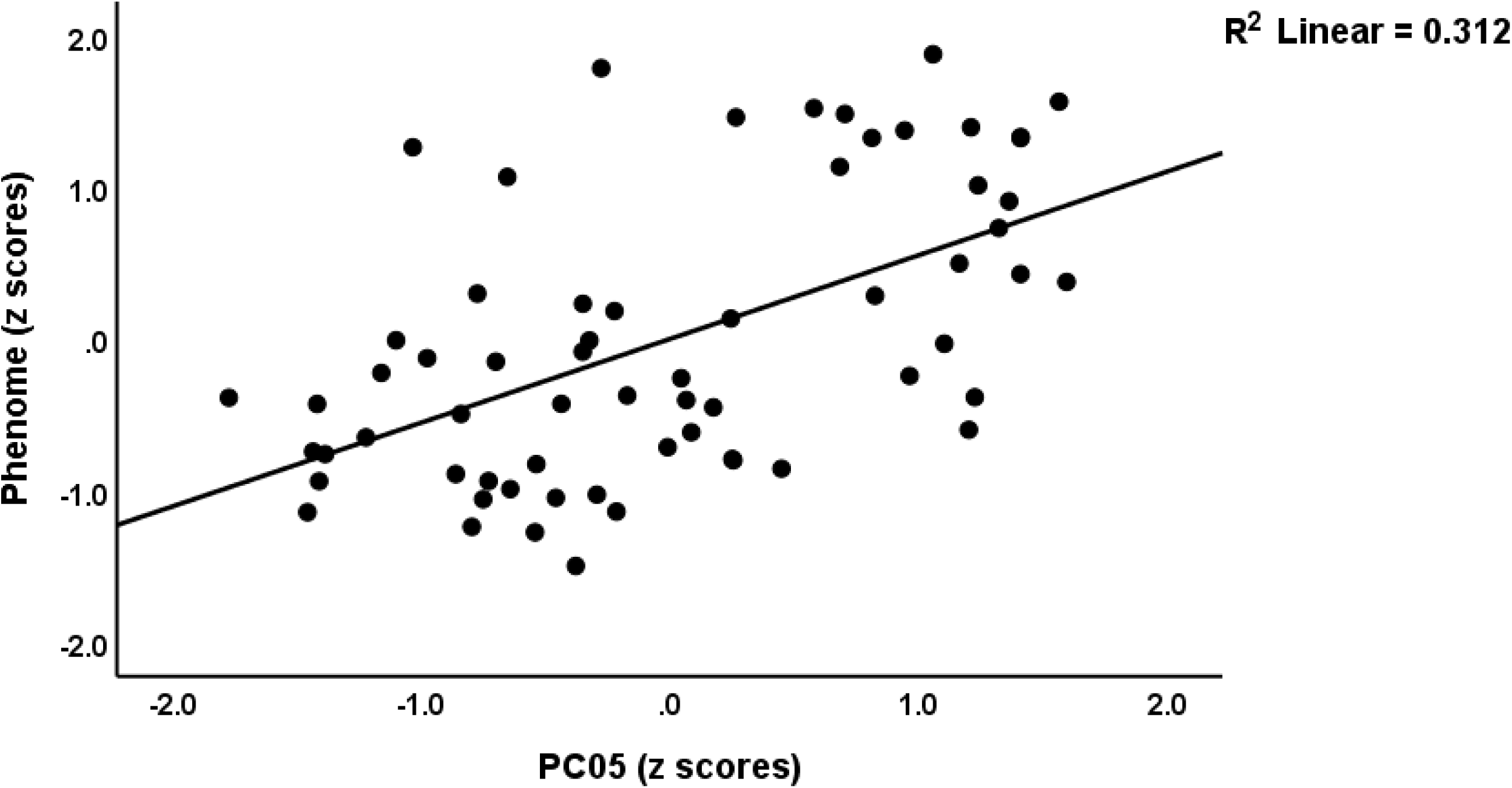
Partial regression of the phenome of maj r depression (MDD) score on the principal compone score PC05, which reflects an upregulated PC network in MDD. See ESF, Figure 4 for the immune data th used to construct this PC (p<0.001).

### Interactions patterns between MDD and MetS

ESF, Table 7 examines the interaction patterns between MDD x MetS using univariate GLM analyses with MDD and MetS and their interaction as fixed factors, while adjusting for the effects of age and sex. This table shows that there were significant interaction effects for TNF signaling, PC04, PC05 and PC06 (all increased in MDD+MetS) and IL-1 signaling and PCinv (both lowered in MDD+MetS). These interaction patterns are shown in ESF, Figures 1-6. ESF, Table 8 shows the results of the univariate GLM analyses conducted on cytokines indicating significant interaction patterns between MDD and MetS. ESF, Figures 7-21 display the cytokines that showed significant interaction effects, namely sIL-1RA, IL-4, IL-17, CCL2, PDGF, CCL5, TNF-α and TNF-β (all increased in MDD+MetS), and FGF, IFN-2α, IL-13, and VEGF (all decreased in MDD+MetS). All in all, while there are no significant differences in any of the cytokines between people with and without MetS, effects of MetS appeared in patients with MDD.

Given that there are significant interactions between MDD and MetS, we have also examined the correlations between the features of MDD and the PCs and their interactions with MetS (see **Table 6**). We did not find any correlations between PC04 and the clinical MDD features, whereas the latter were significantly and positively correlated with PC04 x MetS. The correlation coefficients between PC05 x MetS were greater than those obtained for PC05 alone. In contrast, there were no significant associations between the clinical features and either MetS or BMI.

Given that there are significant interactions between MDD and MetS we have also rerun the regression analyses by entering interaction terms between immune data (including PC05 and PCinv and the cytokines that showed significant interactions when considering MDD and MetS, e.g. CCL5, TNF-β, VEGF, TNF signaling, etc). Table 2 shows that, when using the total study group, the combination of these interaction terms (e.g. PC05 x MetS, PCinv x MetS) significantly predicted MDD with a Nagelkerke value of 0.373 (X^2^=43.704, df=2, p<0.001). The same table shows that using CCL5 x MetS, TNF-β x MetS, VEGF x MetS, combined with CCL11, and M-CSF yielded a good accuracy (82.7%) with a sensitivity of 84.8%, specificity 80.6% (Nagelkerke=0.613 X^2^=81.962, df=5, p<0.001). The interactions between the immune data X BMI always showed a less favorable outcome than the interactions between immune data x MetS.

We reran the multiple regression analyses shown in Table 3 (all subjects combined) by entering the abovementioned interaction terms. These results (regressions #7 - #9) show that we were able to improve the predictions of ROI, neuroticism, suicidal behaviors and the phenome by entering interaction terms including CCL5 x MetS and PCinv x MetS (ROI: R^2^ improved from 0.276 to 0.390), TNF-β x MetS (neuroticism: R^2^ improved from 0.155 to 0.266), CCL5 x MetS and VEGF x MetS (suicidal behaviors: R^2^ improved from 0.102 to 0.236), and TNF signaling x MetS and CCL5 x MetS (phenome: R^2^ improved from 0.265 to 0.314).

### PPI analysis

Tables 9-13 of the ESF show the outcome of PPI analyses performed on the upregulated and downregulated DEPs. **Figure 4** displays that the 20 PC05 DEPs shape a tight PPI network. The PPI plot shows the DEPs that were enriched in “regulation of response to external stimulus” and “defense response”. Figure 5 shows that the 10 DEPs, which were upregulated in MDD, shaped a tight network that was strongly associated with 4 GO annotations, namely “immune response”, “defense response”, “response to stress”, and “inflammatory response”.

**Figure 4.**
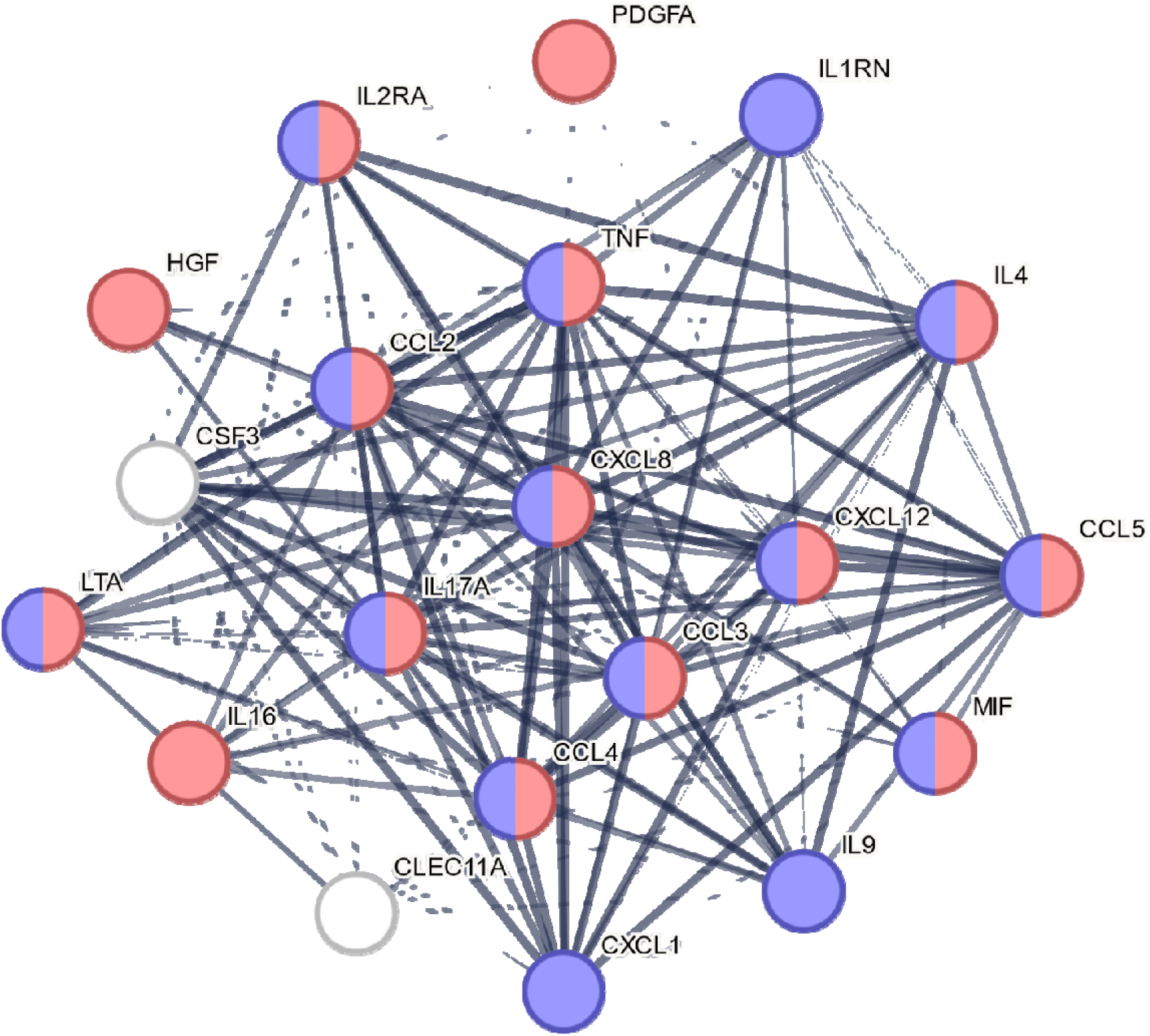
Results of protein-protein interaction (PI) analysis with the zero order PPI network based on an upregulated cytokine, chemokine and growth factor network in major depression. The network features are: 20 nodes, 164 edges, average node degree=16.4, average local clustering coefficient=0.913, expected number of edges=19, PPI enrichment value <1e-16. See ESF, Table 1 for explanation of the differentially expressed proteins, and ESF, Table 4 for computation of the network based on principal component analysis (PC05). Selected top GO enriched terms are shown in colors, red: regulation of response to external stimulus; pink: defense response.

**Figure 5.**
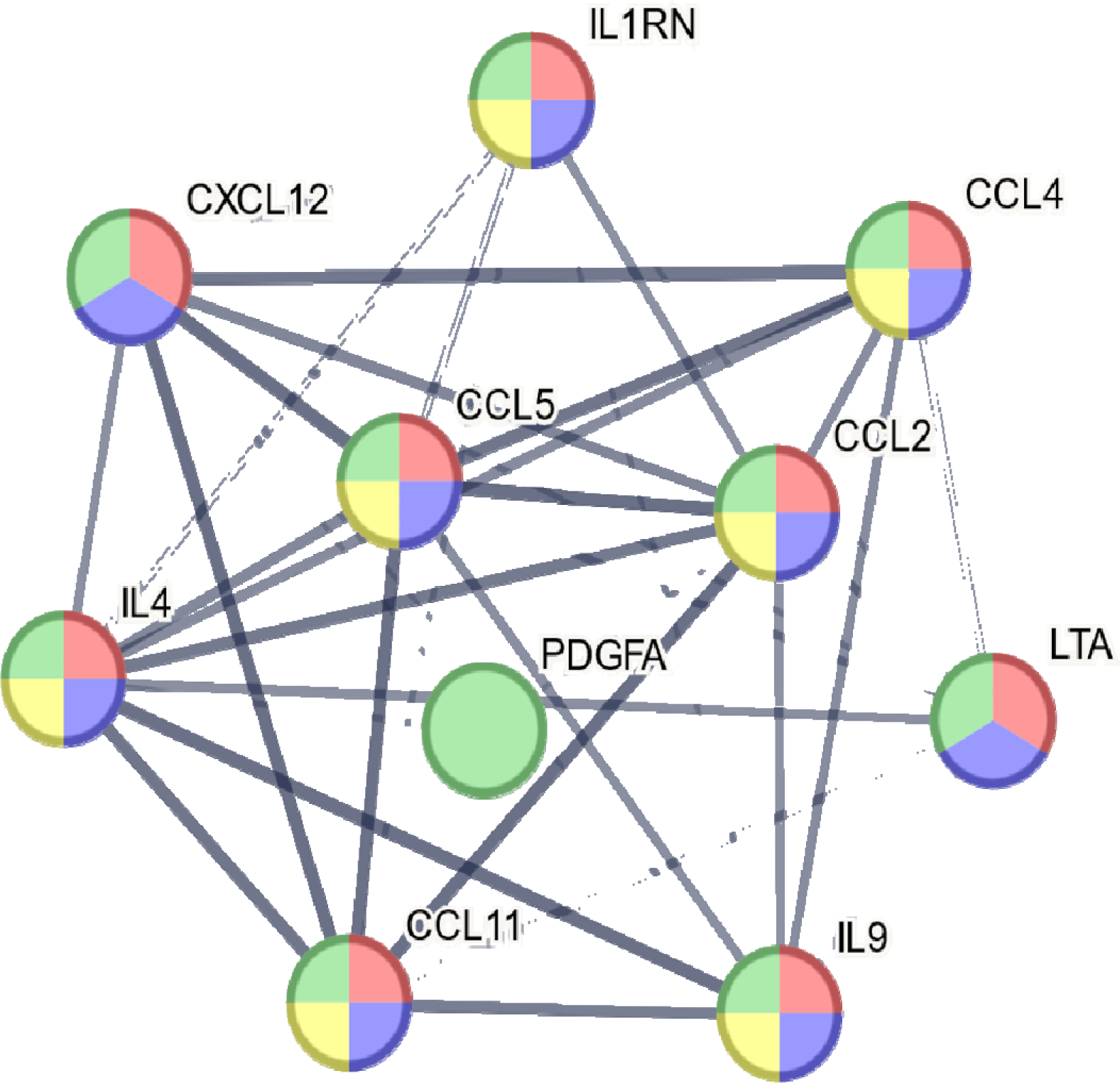
Results of protein-protein interaction (PPI) analysis showing the zero order PPI network based on the differentially expressed proteins (DEPs) that were upregulated in major depression. The network features are: 10 nodes, 38 edges, av rage node degree=7.6, average local clustering coefficient=0.927, expected number of edges=3, PPI enrichment value <1e-16. See Figure 1 and ESF, table 1 for explanation of the DEPs. Selected top GO enriched terms are shown in colors; red: immune response; blue: defense response; green: response to stress; yellow: inflammatory response.

## Discussion

### Immune profiles of the MDD outpatient sample

The first major finding of this research is that OMDD patients do not exhibit any of the conventional immune profiles (M1, alternative M2, Th-1, Th-2, IRS, CIRS) that were identified in mild FE-OMDD (reduced CIRS), severe IMDD (T cell activation and inflammatory profile) or severe FE-IMDD (Th-2, Th-17, IRS, and CIRS) patients. On the contrary, we identified a distinct profile in OMDD by examining alterations in two tightly connected immune networks. Elevated concentrations of chemokines (CXCL12, CCL2, CCL4, CCL11), cytokines (TNF-β and TNF signaling, IL-4, IL-9, and sIL-1RA), and growth factors (PDGF) characterize the first profile. Reduced concentrations of growth factors (NGF, VEGF, M-CSF), cytokines (IL-10, IL-1β, and IL-12p40), and one chemokine (CCL3) characterize the second.

Comparing results with previous publications is challenging since most prior research on MDD has only measured a small number of cytokines, primarily M1 cytokines (e.g., TNF-α and IL-6), in addition to IL-10, sIL-2R, CXCL8, and occasionally other chemokines. Furthermore, none of the papers authored by other researchers computed specific immune profiles in specific MDD phenotypes. In contrast, the present study, Maes et al. (Michael, Asara et al. 2023), and Almulla et al. (Almulla, Ali Abbas Abo et al. 2023a) utilized comprehensive immune profiles constructed from 48 cytokine assays in different MDD phenotypes. Furthermore, all three studies’ assays were conducted simultaneously (over the course of two consecutive days) in the same laboratory, utilizing identical kits (BioRad 48-plex) and under the guidance of the same operator. Consequently, it is possible to compare the outcomes of these three investigations conducted on unique phenotypes of MDD.

The immune profiles of MDD observed in the present investigation diverges from the profiles previously identified in mild FE-OMDD (Michael, Asara et al. 2023) and severe FE-IMDD (Almulla, Ali Abbas Abo et al. 2023a). A significant reduction in the CIRS profile was observed in the former because of decreased levels of CIRS cytokines and M2 and Th2 profiles (Michael, Asara et al. 2023). Conversely, Almulla et al. (Almulla, Ali Abbas Abo et al. 2023a) identified activated Th-1, Th-2, and Th-17 profiles in severe FE-IMDD, along with Th-1 polarization.

Immunological distinctions among other discrete subtypes of MDD have been previously established. Maes et al. (Maes, Rachayon et al. 2022) identified substantial distinctions between mild and very severe MDD through stimulated (LPS+PHA) whole blood assays. Specifically, individuals with mild MDD did not demonstrate changes in IRS activation, M1, Th-1, Th-17, Th-2, CIRS, or growth factor activation, whereas those with severe depression did. According to these results, severe MDD is distinguished by immune system sensitization, including sensitization of the IRS and CIRS pathways. Additionally, notable distinctions were identified based on the stage of the index episode: the acute phase is distinguished by intense IRS activation, whereas partial remission is characterized by predominance of CIRS activities, which indicate a state of repair (Maes and Carvalho 2018). To illustrate, bipolar depressed patients exhibit heightened T cell activation and effector cells during the acute phase, while there is a substantial increase in Treg cells during the partial remission phase (Maes, Nani et al. 2021).

### Cytokines/chemokines/growth hormones in subacute MDD in outpatients

The second major finding of this study is that for OMDD patients, CXCL12, TNF-β, PDGF (all increased), and VEGF (decreased) were the most significant indicators, whilst also M-CSF contributed to the prediction of OMDD. On the other hand, Maes et al. (Michael, Asara et al. 2023) identified IL-4, G-CSF, and CCL11 as the top three predictors of mild FE-OMDD. PDGF and IL-16 were the most accurate predictors of depression symptoms in severe FE-IMDD (Almulla, Ali Abbas Abo et al. 2023a). Meta-analyses conducted on MDD have revealed that elevated levels of TNF-α and IL-6 are associated with this disorder (Köhler, Freitas et al. 2017). In our study, OMDD is indeed characterized by elevated TNF signaling; however, elevated TNF-β was more significant than elevated TRAIL and TNF-α. Almulla et al. (Almulla, Ali Abbas Abo et al. 2023a) demonstrated that severe FE-IMDD patients are distinguished not only by elevated TNF-β, but particularly by elevated TRAIL while TNF-α was only moderately increased. CXCL12, TNF-β, PDGF, CCL11, IL-9, IL-4, CCL5, CCL2, CCL4, and sIL-1RA were among the additional cytokines that increased in the current investigation. Almulla et al. (Almulla, Ali Abbas Abo et al. 2023a) identified elevated levels of sIL-1RA, sIL-2R, IL-6, IL-9, IL-16, IL-18, TNF-α, TNF-β, TRAIL, CCL2, CCL4, CCL5, CCL11, CXCL27, CXCL10, CXCL12, SCGF, PDGF, and HGF in severe FE-IMDD. Thus, both the current cohort and Almulla’s sample are, therefore, distinguished by elevated concentrations of sIL-1RA, CXCL12, TNF-β, PDGF, CCL11, IL-9, CCL2, CCL4, and CCL5.

A recent meta-analysis (Köhler, Freitas et al. 2017) found that the concentration of the master negative immunoregulatory cytokine IL-10 was significantly higher in MDD patients (phenotypes not specified) than in controls; however, in the current study group and the (Michael, Asara et al. 2023) study, this cytokine was significantly lower than in controls. Almulla et al. (Almulla, Ali Abbas Abo et al. 2023a) were unable to identify a substantial elevation in IL-10 levels among FE-IMDD patients, suggesting that this cytokine is relatively deficient in relation to the substantially increased Th-1 polarization and IRS activation. In the study published by Almulla et al. (Almulla, Ali Abbas Abo et al. 2023a), VEGF levels were also reduced; however, in cases of mild FE-OMDD, VEGF levels remained unchanged (Michael, Asara et al. 2023). Maes et al. (Michael, Asara et al. 2023) and the current study both reported decreased M-CSF levels in MDD, whereas Almulla et al. (Almulla, Ali Abbas Abo et al. 2023a) observed substantially elevated M-CSF levels in their study sample. In a cohort from a previous MDD study (Maes, Rachayon et al. 2023), NGF levels were substantially reduced, whereas no significant alterations were observed in mild FE-OMDD (Michael, Asara et al. 2023). Like the present investigation, Almulla et al. (Almulla, Ali Abbas Abo et al. 2023a) identified decreased CCL3 in patients with severe FE-IMDD. IL-4 was significantly decreased in mild FE-OMDD, whereas this cytokine was substantially increased in the current study and in the study sample of Almulla et al. (Almulla, Ali Abbas Abo et al. 2023a). Bipolar patients may have elevated IL-4 levels, according to recent meta-analyses (Modabbernia, Taslimi et al. 2013, Munkholm, Braüner et al. 2013). However, significant parallels exist among the three phenotypes of MDD, which will be elaborated upon in the following section.

### Annotations of the immune profiles in OMDD

The upregulated and downregulated immune DEP networks in the current OMDD study group reflect distinct functions, according to our PPI findings. First, the tight network of the upregulated DEPs was found to be significantly enriched in a “defense response”, “immune response”, “regulation of response to external stimulus”, “response to stress”, “inflammatory response”, and “positive regulation of a response to external stimulus”. Furthermore, the PPI analysis demonstrates that the pathways that were enriched in the networks supporting the downregulated DEPs were “chemotaxis”, “positive regulation of neurogenesis”, “negative regulation of neuronal death, and cell apoptotic process and programmed cell death”, and “positive regulation of cell communication”. Highly significant associations were found between a subset of these downregulated DEPs (CCL3, CSF1, VEGFA, NGF, and IL10) and the “regulation of neuron death” and “positive regulation of neurogenesis”.

Significantly, the DEP network that is upregulated and comprises CCL11, CCL5, CCL2, CCL4, CXCL12, TNF-β, TNF signaling, IL-4 and IL-9 conveys a profound neurotoxic profile. CXCL12 is generated by glial cells and neurons in various regions of the brain (Guyon 2014, Guyon 2014). By increasing p38 MAPK activity, this chemokine is neurotoxic in neuronal-glial cerebrocortical cell cultures (Sanchez, Medders et al. 2016). Moreover, the CXCL12 system exhibits numerous cross-talks with glutamate, GABA, cannabinoids, and opioids (Guyon 2014, Guyon 2014). It has been previously mentioned that heightened TNF signaling can lead to various neurotoxic consequences (Maes and Carvalho 2018, Maes, Sirivichayakul et al. 2020, Almulla, Ali Abbas Abo et al. 2023a). These effects can manifest as cell death and glutamate release (van Loo and Bertrand 2023). TNF-β, which is a pro-inflammatory cytokine, is significant in neurodegenerative and cytotoxic processes (James Bates, Browne et al. 2022). CCL11 can cross the blood-brain barrier (BBB) (Erickson, Morofuji et al. 2014) and inhibits neurogenesis, disrupts the blood-brain barrier, and induces neurotoxicity via an increase in oxidative stress (Jamaluddin, Wang et al. 2009, Parajuli, Horiuchi et al. 2015, Ivanovska, Abdi et al. 2020). However, prior research on the role of CCL11 in MDD produced contradictory results (Leighton, Nerurkar et al. 2018, Teixeira, Gama et al. 2018, Al-Hakeim, Najm et al. 2020, Almulla, Ali Abbas Abo et al. 2023a). CCL2 has the potential to cross the BBB and disrupt the tight junctions of the BBB, which could result in an increase in blood flow across the BBB (Dimitrijevic, Stamatovic et al. 2006, Guyon, Skrzydelski et al. 2009). CCL2 has the potential to induce neurotoxicity by modulating neuroinflammation via its binding to CCR2 and by stimulating T lymphocytes, macrophages, and dendritic cells; thus, it enhances the behavioral or neurotoxic effects of all cytokines generated by these cell lines (Guyon, Skrzydelski et al. 2009, Kiyota, Gendelman et al. 2013, Zhang and Luo 2019). Elevated plasma concentrations of CCL2 were observed in depressed patients relative to the control group, according to a recent meta-analysis (Leighton, Nerurkar et al. 2018). Astrocytic, microglial, and endothelial cells secrete CCL2 and CCL5 in response to lesions, as well as expressing their chemokine receptors (Glabinski, Tani et al. 1997). In Parkinson’s disease patients, dopaminergic neurons in the substantia nigra may be lost because of CCL5, which also causes deficits in motor functions (Dutta, Kundu et al. 2019). Patients diagnosed with menstrual cycle associated syndrome, characterized by symptoms including anxiety, depression, and fatigue, exhibit elevated levels of both CCL2 and CCL5 in their serum (Roomruangwong, Sirivichayakul et al. 2020). CCL4, a chemokine with pro-inflammatory properties, has the potential to induce BBB breakdown (Estevao, Bowers et al. 2021). IL-4 may increase neurotoxicity in the cortex via oxidative stress (Jeong, Chung et al. 2019). IL-9, which may be produced by Th cells in the presence of IL-4, may promote Th-17 migration into the brain (Zhou, Sonobe et al. 2011). Furthermore, PDGF, which by itself displays neuroprotective effects, may activate the cytokine-chemokine network (Maes, Plaimas et al. 2021).

Based on the upregulation of DEPs that induce BBB breakdown and exert neurotoxic effects, and the downregulation of DEPs that positively regulate neurogenesis and negatively regulate neuron death, it can be inferred that OMDD patients exhibit a severe neurotoxic immune profile. Consequently, severe FE-IMDD and the current study population are comparable in that they both involve heightened immunoneurotoxicity. In FE-OMDD, an elevated concentration of the neurotoxic CCL11 and G-CSF, in addition to a diminished CIRS, predicted the syndrome manifestation (Michael, Asara et al. 2023). While G-CSF exhibits neuroprotective properties over an extended period, it may impair inflammation and neutrophil-mediated immunity (Song, Joe et al. 2016, Martin, Wong et al. 2021).

The elevation in IL-4 levels observed in the current sample can likely be attributed to the immune system’s endeavors to counteract the Th-1 polarization and T cell activation that were prevalent in severe FE-IMDD. An additional CIRS component, sIL-1RA, was found to be elevated in both the current and severe FE-IMDD but remained unchanged in mild FE-OMDD. However, IL-10, an additional important regulator of Treg functions, was diminished in both OMDD and mild FE-OMDD, whereas patients with severe FE-IMDD exhibited a relative deficiency of IL-10 (Almulla, Ali Abbas Abo et al. 2023a). Consequently, decreased IL-10 appears to be the primary drug target in all three subtypes of MDD, because lowered IL-10 may potentially elevate the risk of immunoneurotoxicity and Th-1 responses.

Most importantly, based on the above we may conclude that the features of OMDD, including neuroticism, cognitive symptoms of depression, suicidal behaviors, and the phenome of OMDD are at least in part mediated via the immunoneurotoxic effects of different cytokines, chemokine, and growth factors. Previously, it was shown that in different MDD phenotypes, suicidal behaviors and the pure cognitive symptoms of depression, suicidal ideation and attempts were significantly associated with neuro-immune pathways (Maes, Rachayon et al. 2022, Vasupanrajit, Jirakran et al. 2022, Maes, Rachayon et al. 2023).

### Effects of MetS on immune profiles

After controlling for the influence of MDD (as well as age and sex), the levels of cytokines, chemokines, and growth factors did not differ significantly between subjects with and without MetS, which constitutes the third main finding of this investigation. Previous assertions that MetS is accompanied by inflammatory indicators, as delineated in the Introduction (Di Lorenzo, Dell’agli et al. 2013, Zafar, Khaliq et al. 2018, Chan, Cathomas et al. 2019, Reddy, Lent-Schochet et al. 2019), are not corroborated by these findings. The findings of our study do not support the hypothesis put forth by Chan et al. (Chan, Cathomas et al. 2019) that inflammation serves as a common pathway connecting MDD and MetS. Nonetheless, we discovered that BMI was substantially and positively associated with TRAIL, G-CSF, HGF, sIL-1RA, sIL-2R, IL-16, IL-18, CXCL10, LIF, M-CSF, CCL3, and TRAIL, albeit with a small effect size. Thus, an increase in BMI appears to be associated with a modest upregulation of immune functions, but not MetS per se.

Significant interactions between OMDD and MetS induce substantial increases in TNF signaling, upregulation of key cytokines including IL-4, IL-17, TNF-α, CCL2, CCL5, and PDGF, and upregulation of the DEP neurotoxic networks (PC04, PC05, and PC06). Furthermore, the substantial interactions that occurred between OMDD and MetS resulted in an additional substantial inhibition of the downregulated DEP (PCinv) as well as crucial protective growth factors including VEGF and FGF. MetS therefore contributes to an increase in immunoneurotoxicity in OMDD. Prior research has indicated that increased atherogenicity indices, which are linked to oxidative and nitrosative stress, are present when MDD and MetS are co-occurring (Nunes, Piccoli de Melo et al. 2015, de Melo, Nunes et al. 2017, Morelli, Maes et al. 2021).

Consistent with the aforementioned results, we discovered that the relationships between immune biomarkers and the manifestation of OMDD in individuals with MetS are quite distinct from those without. Consistent with our discovery that the interaction term OMDD X MetS is associated with remarkably significant immune alterations, we observed a stronger correlation between the biomarkers and OMDD features in MetS than in those without MetS. Naturally, this could be accounted for by our discovery that MetS in OMDD elevates immunoneurotoxicity. Significantly, we observed that interaction terms between MetS and key cytokines, including CCL5, CXCL12, TNF-β, VEGF, as well as upregulated and downregulated immune networks, significantly enhance the prediction of the phenome of OMDD in subjects lacking MetS.

## Limitations

The inclusion of oxidative and nitrosative stress markers, which are linked to MetS (Nunes, Piccoli de Melo et al. 2015, de Melo, Nunes et al. 2017, Morelli, Maes et al. 2021), would have enhanced the overall appeal of this paper. Further investigation is warranted into the transitional alterations that occur in the immune networks of inpatients undergoing extremely severe depression as they transition from the acute phase to the subacute state once they are discharged. Additionally, prospective studies should investigate the distinctions between the remitted, partial remitted, and non-response phases of the immune profile measured here. Antidepressants were given to some of the outpatients. Despite these drugs’ significant in vitro effects on M1 and Th-1 cytokine production (lowered) and IL-10 production (increased) (Xia, DePierre et al. 1996, Maes, Song et al. 1999), it is unlikely that they will have many in vivo effects. This is likely because the immune system’s sensitization has not been resolved and etiological factors are still present (Maes, Berk et al. 2012, Maes, Vojdani et al. 2021). Accordingly, the neuro-immune indicators assessed in the current study, were not affected by the medication status. In addition, our results show that IL-10 is still very lower in OMDD patients despite that some patients were treated with antidepressants, which increase IL-10 production.

## Conclusions

OMDD is associated with upregulation of DEPs which indicate the presence of an immune, defense, or stress response. The downregulation of six other DEPs suggests that neurogenesis and regulation of neuron death processes may be diminished. Consequently, OMDD is characterized by increased immunoneurotoxicity. Prominent interactions between OMDD and MetS result in substantial increases in TNF signaling, upregulation of neurotoxic DEPs and downregulation of generally more protective DEPs.

The MDD phenome, suicidal tendencies, and neuroticism were all significantly predicted by increased neurotoxic immune profiles and lowered protective profiles and interactions among those profiles and MetS, causing exaggerated immunoneurotoxicity. The latter profile contributes significantly to neuroticism, suicidal behaviors, and the severity of the phenome of OMDD.

Although there are notable distinctions in the immune profiles of OMDD, mild FE-OMDD, and severe FE-IMDD, all three conditions share a characteristic feature of reduced IL-10 levels. Thus, IL-10 represents a novel therapeutic target for various phenotypes of MDD. Future immune research on MDD should consistently consider the effects of MetS and the variations between distinct MDD phenotypes. The use of antidepressants may be linked to an increased susceptibility to MetS (Gramaglia, Gambaro et al. 2018). Therefore, further inquiry is necessary to thoroughly examine the putative in vivo effects of antidepressants on immune functions via moderating or mediating effects of the MetS.

## Author’s contributions

Conceptualization and study design: MM and JK; first draft writing: MM; statistical analysis: MM; editing: all authors; recruitment of patients: JK. All authors approved the final version of the manuscript.

## Ethics approval and consent to participate

The research project (#445/63) was approved by the Institutional Review Board of Chulalongkorn University’s institutional ethics board. All patients and controls gave written informed consent prior to participation in the study.

## Data Availability Statement

The dataset generated during and/or analyzed during the current study will be available from the corresponding author upon reasonable request and once the dataset has been fully exploited by the authors.

