## supplementary file for "Major depressive disorder, neuroticism, suicidal behaviors, and depression severity are all associated with neurotoxic immune networks and their intricate interactions with metabolic syndrome"

**ELECTRONIC SUPPLEMENTARY FILE (ESF)**

**ESF, Table 1**. Overview of the cytokines, chemokines, and growth factors measured in the current study

| **Protein abbreviations** | **Gene Symbol** | **> OOR (%)** | **Protein name / alias** |
| --- | --- | --- | --- |
| **IFN-α2** | **IFNA2** | 60.8 | Interferon-α2 |
| **IFN-γ** | **IFNG** | 81.9 | Interferon-γ |
| **IL-1α** | **IL1A** | 60.8 | Interleukin-1α |
| **IL-1β** | **IL1B** | 78.3 | Interleukin-1β |
| **sIL-1RA** | **IL1RN** | 100 | Soluble interleukin-1 receptor antagonist |
| **IL-2** | **IL2** | 24.6 | Interleukin-2 |
| **IL-2R** | **IL2RA** | 100 | Soluble interleukin-2 receptor |
| **IL-3** | **IL3** | 0 | Interleukin-3 |
| **IL-4** | **IL4** | 100 | Interleukin-4 |
| **IL-5** | **IL5** | 5.8 | Interleukin-5 |
| **IL-6** | **IL6** | 27.5 | Interleukin-6 |
| **IL-7** | **IL7** | 2.9 | Interleukin-7 |
| **IL-9** | **IL9** | 100 | Interleukin-9 |
| **IL-10** | **IL10** | 56.4 | Interleukin-10 |
| **IL-12p70** | **IL12A/B** | 37.0 | Interleukin-12 p70 |
| **IL-12p40** | **IL12B** | 60.8 | Interleukin-12 p40 |
| **IL-13** | **IL13** | 68.1 | Interleukin-13 |
| **IL-15** | **IL15** | 10.9 | Interleukin-15 |
| **IL-16** | **IL16** | 100 | Interleukin-16 |
| **IL-17** | **IL17A** | 60.8 | Interleukin-17 |
| **IL-18** | **IL18** | 100 | Interleukin-18 |
| **TNF-α** | **TNF** | 100 | Tumor necrosis factor-α |
| **TNF-β** | **LTA** | 100 | Tumor necrosis factor-β or lymphotoxin-alpha (LT-α) |
| **TRAIL** | **TNFSF10** | 100 | TNF-related apoptosis-inducing ligand (TRAIL) or tumor necrosis factor ligand superfamily member 10 (TNFSF10) |
| **LIF** | **LIF** | 94.2 | Leukemia inhibitory factor |
| **MIF** | **MIF** | 100 | Macrophage migration inhibitory factor-like protein (MIF) or glycosylation-inhibiting factor |
| **G-CSF** | **CSF3** | 100 | Granulocyte colony stimulating factor (G-CSF) or colony stimulating factor 3 (CSF3) |
| **M-CSF** | **CSF1** | 100 | Macrophage colony-stimulating factor (M-CSF) or colony stimulating factor 1 (CSF1) |
| **GM-CSF** | **CSF2** | 60.8 | Granulocyte-macrophage colony-stimulating factor (GM-CSF) or colony-stimulating factor 2 (CSF2) |
| **CCL2 or MCP1** | **CCL2** | 100 | C-C motif chemokine ligand 2 (CCL2) or monocyte chemoattractant protein 1 (MCP1) |
| **CCL3 or MIP-1α** | **CCL3** | 100 | C-C motif Chemokine ligand 3 (CCL3) or macrophage inflammatory protein 1-alpha (MIP-1α) |
| **CCL4 or MIP-1β** | **CCL4** | 100 | C-C motif chemokine ligand 4 (CCL4) or macrophage inflammatory protein 1β (MIP-1β) or lymphocyte activation gene 1 protein |
| **CCL5 or RANTES** | **CCL5** | 100 | C-C motif chemokine ligand 5 (CCL5) or regulated upon activation, normally T-expressed, and presumably Secreted (RANTES) |
| **CCL7 or MCP3** | **CCL7** | 60.8 | C-C motif chemokine ligand 7 (CCL7) or monocyte-chemotactic protein 3 (MCP3). |
| **CCL11 or Eotaxin** | **CCL11** | 100 | C-C motif chemokine ligand 11 (CCL11) or eosinophil chemotactic protein |
| **CCL27 or CTACK** | **CCL27** | 100 | C-C motif chemokine ligand 27 (CCL27) or cutaneous T-cell attracting chemokine (CTACK) |
| **CXCL1 or GRO-α** | **CXCL1** | 100 | C-X-C motif chemokine 1 (CXCL1) or growth-regulated alpha protein (GRO) |
| **CXCL8 or IL-8** | **CXCL8** | 100 | C-X-C motif chemokine ligand 8 (CXCL8) or interleukin-8 (IL-8) |
| **CXCL9 or MIG** | **CXCL9** | 100 | C-X-C motif chemokine ligand 9 (CXCL9) or monokine induced by gamma interferon (MIG) |
| **CXCL10 or IP10** | **CXCL10** | 100 | C-X-C motif chemokine ligand 10 (CXCL10) or Interferon gamma-induced protein 10 (IP10) |
| **CXCL12 or SDF-1α** | **CXCL12** | 100 | C-X-C motif chemokine 12 (CXCL12) or stromal cell-derived factor 1 (SDF-1α) |
| **FGF** | **FGF2** | 91.3 | Fibroblast growth factor 2 (FGF) or basic fibroblast growth factor |
| **HGF or SF** | **HGF** | 100 | Hepatocyte growth factor (HGF) or scatter factor (SF) |
| **BNGF** | **NGF** | 63.8 | β-nerve growth factor (NGF) |
| **PDGF** | **PDGFA** | 100 | Platelet derived growth factor (PDGF) |
| **SCF** | **KITLG** | 100 | Stem cell factor (SCF) or Kit ligand (KITLG) |
| **SCGF-β or CLEC11A** | **CLEC11A** | 100 | Stem cell growth factor (SCGF) or C-type lectin domain family 11 member A (CLEC11A) |
| **VEGF** | **VEGFA** | 89.1 | Vascular endothelial growth factor (VEGF) |

Adapted from: *Maes M, Rachayon M, Jirakran K, Sodsai P, Klinchanhom S, Gałecki P, Sughondhabirom A, Basta-Kaim A. The Immune Profile of Major Dysmood Disorder: Proof of Concept and Mechanism Using the Precision Nomothetic Psychiatry Approach. Cells. 2022 Mar 31;11(7):1183. doi: 10.3390/cells11071183. PMID: 35406747; PMCID: PMC8997660.*

*Kalayasiri R, Dadwat K, Supaksorn T, ,Sirivichayakul S, Maes M. Methamphetamine (MA) use, MA dependence, and MA-induced psychosis are associated with increasing aberrations in the compensatory immunoregulatory system and interleukin-1α and CCL5 levels. medRxiv 2023.03.26.23287766; doi: https://doi.org/10.1101/2023.03.26.23287766*

**ESF, Table 2. Description of the immune profiles used in this study**

| **Immune Profile** | **Members** |
| --- | --- |
| **M1 macrophage** | IL-1β, IL-6, TNF-α, IL-12p70, IL-15, CCL2, CCL5, CXCL1, CXCL8, CXCL9, CXCL10 |
| **M2 macrophages** | IL-10, IL-4, IL-13, VEGF, PDGF, sIL-1RA |
| **z M1 – z M2** | zM1 – zM2 |
| **T helper (Th)-1** | IL-2, sIL-2R, IFN-a, IFN-γ, IL-12p70, IL-16, TNF-α, TNF-β |
| **Th-2** | IL-4, IL-5, IL-9, IL-13, IL-10, IL-6 |
| **z Th- z Th-2** | zTh-1 – zTh-2 |
| **IRS** | IL-1α, IL-1β, IL-6, IL-12p70, IL-15, IL-16, IL-17, IL-18, CCL2, CCL3, CCL4, CCL5, CCL7, CCL11, CXCL1, CXCL8, CXCL9, CXCL10, IL-2, IFN-α, IFN-γ, TNF-α, TNF-β, TRAIL, GM-CSF, M-CSF, G-CSF, SCGF |
| **CIRS** | IL-4, IL-10, sIL-1RA, sIL-2R |
| **z IRS – z CIRS** | zIRS – zCIRS |
| **IL-1 signaling** | aIL-1α + zIL-1β - zsIL1RA |
| **TNF signaling** | TNF-α, TNF-β, TRAIL |

IRS: immune-inflammatory response system; CIRS: compensatory immunoregulatory system

Adapted from:

*Maes M, Rachayon M, Jirakran K, Sodsai P, Klinchanhom S, Gałecki P, Sughondhabirom A, Basta-Kaim A. The Immune Profile of Major Dysmood Disorder: Proof of Concept and Mechanism Using the Precision Nomothetic Psychiatry Approach. Cells. 2022 Mar 31;11(7):1183. doi: 10.3390/cells11071183. PMID: 35406747; PMCID: PMC8997660.*

*Kalayasiri R, Dadwat K, Supaksorn T, ,Sirivichayakul S, Maes M. Methamphetamine (MA) use, MA dependence, and MA-induced psychosis are associated with increasing aberrations in the compensatory immunoregulatory system and interleukin-1α and CCL5 levels. medRxiv 2023.03.26.23287766; doi: https://doi.org/10.1101/2023.03.26.23287766*

**ESF, Table 3a. Differences in immune profiles between patients with major depression (MDD, 1) and healthy controls (noMDD, 0).**

| **Estimates** | | | | | |
| --- | --- | --- | --- | --- | --- |
| Dependent Variable | MDD_NoMDD | Mean | Std. Error | 95% Confidence Interval | |
|  |  |  |  | Lower Bound | Upper Bound |
| M1 | .00 | -.059^a^ | .124 | -.305 | .186 |
|  | 1.00 | .063^a^ | .125 | -.185 | .311 |
| M2 | .00 | -.098^a^ | .121 | -.337 | .142 |
|  | 1.00 | .099^a^ | .122 | -.143 | .341 |
| zM1-zM2 | .00 | .046^a^ | .122 | -.195 | .288 |
|  | 1.00 | -.044^a^ | .123 | -.287 | .200 |
| Th-1 | .00 | -.084^a^ | .124 | -.329 | .161 |
|  | 1.00 | .086^a^ | .125 | -.161 | .334 |
| Th-2 | .00 | -.063^a^ | .123 | -.305 | .180 |
|  | 1.00 | .058^a^ | .124 | -.187 | .303 |
| zTh1-zTh2 | .00 | -.003^a^ | .123 | -.247 | .241 |
|  | 1.00 | .009^a^ | .124 | -.237 | .255 |
| IRS | .00 | -.034^a^ | .125 | -.280 | .213 |
|  | 1.00 | .033^a^ | .126 | -.216 | .282 |
| CIRS | .00 | .042^a^ | .124 | -.203 | .286 |
|  | 1.00 | -.047^a^ | .125 | -.294 | .200 |
| zIRS-zCIRS | .00 | -.088^a^ | .124 | -.333 | .157 |
|  | 1.00 | .093^a^ | .125 | -.154 | .341 |
| IL-1 signaling | .00 | .280^a^ | .112 | .058 | .503 |
|  | 1.00 | -.294^a^ | .114 | -.519 | -.070 |
| TNF signaling | .00 | -.265^a^ | .120 | -.502 | -.028 |
|  | 1.00 | .272^a^ | .121 | .033 | .512 |
| a. Covariates appearing in the model are evaluated at the following values: Sex = .203, Age = 37.436. | | | | | |

All data are shown as z scores.

| **Univariate Tests** | | | | | | |
| --- | --- | --- | --- | --- | --- | --- |
| Dependent Variable | | Sum of Squares | df | Mean Square | F | Sig. |
| M1 | Contrast | .479 | 1 | .479 | .472 | .493 |
|  | Error | 129.867 | 128 | 1.015 |  |  |
| M2 | Contrast | 1.242 | 1 | 1.242 | 1.284 | .259 |
|  | Error | 123.799 | 128 | .967 |  |  |
| zM1-zM2 | Contrast | .261 | 1 | .261 | .266 | .607 |
|  | Error | 125.379 | 128 | .980 |  |  |
| Th-1 | Contrast | .930 | 1 | .930 | .917 | .340 |
|  | Error | 129.770 | 128 | 1.014 |  |  |
| Th-2 | Contrast | .471 | 1 | .471 | .475 | .492 |
|  | Error | 126.982 | 128 | .992 |  |  |
| zTh1-zTh2 | Contrast | .005 | 1 | .005 | .005 | .945 |
|  | Error | 128.143 | 128 | 1.001 |  |  |
| IRS | Contrast | .142 | 1 | .142 | .139 | .710 |
|  | Error | 130.929 | 128 | 1.023 |  |  |
| CIRS | Contrast | .251 | 1 | .251 | .249 | .619 |
|  | Error | 129.029 | 128 | 1.008 |  |  |
| zIRS-zCIRS | Contrast | 1.061 | 1 | 1.061 | 1.049 | .308 |
|  | Error | 129.435 | 128 | 1.011 |  |  |
| IL-1 signaling | Contrast | 10.607 | 1 | 10.607 | 12.733 | .001 |
|  | Error | 106.625 | 128 | .833 |  |  |
| TNF signaling | Contrast | 9.274 | 1 | 9.274 | 9.779 | .002 |
|  | Error | 121.398 | 128 | .948 |  |  |
| All results of univariate GLM analyses with metabolic syndrome as second factor, and age and sex as covariates. The F tests the effect of MDD_NoMDD. This test is based on the linearly independent pairwise comparisons among the estimated marginal means. | | | | | | |

See Table 2 for explanation of the immune profiles.

**ESF, Table 3b. Differences in immune profiles between patients with and without metabolic syndrome (MetS).**

| **Estimates** | | | | | |
| --- | --- | --- | --- | --- | --- |
| Dependent Variable | MetS_NoMetS | Mean | Std. Error | 95% Confidence Interval | |
|  |  |  |  | Lower Bound | Upper Bound |
| M1 | .00 | -.030^a^ | .129 | -.286 | .226 |
|  | 1.00 | .034^a^ | .135 | -.234 | .301 |
| M2 | .00 | .007^a^ | .126 | -.243 | .257 |
|  | 1.00 | -.006^a^ | .132 | -.267 | .255 |
| zM1-zM2 | .00 | -.045^a^ | .127 | -.297 | .207 |
|  | 1.00 | .048^a^ | .133 | -.215 | .310 |
| Th-1 | .00 | -.015^a^ | .129 | -.271 | .242 |
|  | 1.00 | .017^a^ | .135 | -.250 | .284 |
| Th-2 | .00 | .071^a^ | .128 | -.183 | .324 |
|  | 1.00 | -.075^a^ | .134 | -.339 | .189 |
| Th1-Th2 | .00 | -.075^a^ | .129 | -.329 | .179 |
|  | 1.00 | .081^a^ | .134 | -.185 | .347 |
| zIRS | .00 | .018^a^ | .130 | -.239 | .275 |
|  | 1.00 | -.019^a^ | .136 | -.288 | .249 |
| zCIRS | .00 | .054^a^ | .129 | -.201 | .310 |
|  | 1.00 | -.059^a^ | .135 | -.326 | .207 |
| zIRS-zCIRS | .00 | -.042^a^ | .129 | -.298 | .213 |
|  | 1.00 | .047^a^ | .135 | -.220 | .314 |
| IL-1 signaling | .00 | .119^a^ | .117 | -.113 | .351 |
|  | 1.00 | -.133^a^ | .122 | -.375 | .110 |
| TNF signaling | .00 | -.037^a^ | .125 | -.285 | .210 |
|  | 1.00 | .044^a^ | .131 | -.214 | .303 |
| a. Covariates appearing in the model are evaluated at the following values: Sex = .203, Age = 37.436. | | | | | |

| **Pairwise Comparisons** | | | | | | | |
| --- | --- | --- | --- | --- | --- | --- | --- |
| Dependent Variable | (I) MetS_NoMetS | (J) MetS_NoMetS | Mean Difference (I-J) | Std. Error | Sig. | 95% Confidence Interval for Difference^a^ | |
|  |  |  |  |  |  | Lower Bound | Upper Bound |
| M1 | .00 | 1.00 | -.064 | .199 | .748 | -.457 | .329 |
|  | 1.00 | .00 | .064 | .199 | .748 | -.329 | .457 |
| M2 | .00 | 1.00 | .013 | .194 | .947 | -.371 | .396 |
|  | 1.00 | .00 | -.013 | .194 | .947 | -.396 | .371 |
| zM1-zM2 | .00 | 1.00 | -.093 | .195 | .635 | -.479 | .293 |
|  | 1.00 | .00 | .093 | .195 | .635 | -.293 | .479 |
| Th-1 | .00 | 1.00 | -.031 | .199 | .874 | -.424 | .361 |
|  | 1.00 | .00 | .031 | .199 | .874 | -.361 | .424 |
| Th-2 | .00 | 1.00 | .146 | .196 | .460 | -.243 | .534 |
|  | 1.00 | .00 | -.146 | .196 | .460 | -.534 | .243 |
| zTh1-zTh2 | .00 | 1.00 | -.156 | .197 | .430 | -.546 | .234 |
|  | 1.00 | .00 | .156 | .197 | .430 | -.234 | .546 |
| zIRS | .00 | 1.00 | .037 | .199 | .852 | -.357 | .432 |
|  | 1.00 | .00 | -.037 | .199 | .852 | -.432 | .357 |
| zCIRS | .00 | 1.00 | .114 | .198 | .567 | -.278 | .505 |
|  | 1.00 | .00 | -.114 | .198 | .567 | -.505 | .278 |
| zIRS-zCIRS | .00 | 1.00 | -.089 | .198 | .653 | -.482 | .303 |
|  | 1.00 | .00 | .089 | .198 | .653 | -.303 | .482 |
| IL-1 signaling | .00 | 1.00 | .251 | .180 | .165 | -.105 | .607 |
|  | 1.00 | .00 | -.251 | .180 | .165 | -.607 | .105 |
| TNF signaling | .00 | 1.00 | -.082 | .192 | .671 | -.462 | .298 |
|  | 1.00 | .00 | .082 | .192 | .671 | -.298 | .462 |
| All results of univariate GLM analyses with metabolic syndrome as second factor, and age and sex as covariates. The F tests the effect of MDD_NoMDD. This test is based on the linearly independent pairwise comparisons among the estimated marginal means. See Table 2 for explanation of the immune profiles. | | | | | | | |

**ESF, Table 4. Construction four principal components**

**Construction of PC04**

| **KMO and Bartlett's Test** | | |
| --- | --- | --- |
| Kaiser-Meyer-Olkin Measure of Sampling Adequacy. | | .860 |
| Bartlett's Test of Sphericity | Approx. Chi-Square | 2302.773 |
|  | df | 351 |
|  | Sig. | .000 |

| **Component Matrix^a^** | |
| --- | --- |
|  | Component |
|  | 1 |
| CCL27 | .418 |
| FGF | .422 |
| G-CSF | .614 |
| CXCL1 | .510 |
| HGF | .606 |
| IFN-g | .437 |
| sIL-1RA | .658 |
| sIL-2R | .619 |
| IL-4 | .758 |
| IL-8 | .561 |
| IL-9 | .643 |
| IL-16 | .619 |
| IL-17 | .614 |
| CCL2 | .568 |
| M-CSF | .417 |
| MIF | .579 |
| CXCL9 | .416 |
| CCL3 | .550 |
| CCL4 | .744 |
| PDGF | .677 |
| CCL5 | .714 |
| SCF | .424 |
| SCGF | .598 |
| CXCL12 | .575 |
| TNF-a | .731 |
| TNF-b | .618 |
| TRAIL | .432 |
| Extraction Method: Principal Component Analysis. | |
| a. 1 components extracted. | |

**Construction of PC05**

| **KMO and Bartlett's Test** | | |
| --- | --- | --- |
| Kaiser-Meyer-Olkin Measure of Sampling Adequacy. | | .867 |
| Bartlett's Test of Sphericity | Approx. Chi-Square | 1829.627 |
|  | df | 190 |
|  | Sig. | .000 |

| **Component Matrix^a^** | |
| --- | --- |
|  | Component |
|  | 1 |
| G-CSF | .588 |
| CXCL1 | .524 |
| HGF | .625 |
| sIL-1RA | .652 |
| sIL-2R | .568 |
| IL-4 | .756 |
| IL-8 | .556 |
| IL-9 | .684 |
| IL-16 | .602 |
| IL-17 | .607 |
| CCL2 | .560 |
| MIF | .571 |
| CCL3 | .524 |
| CCL4 | .766 |
| PDGF | .718 |
| CCL5 | .740 |
| SCGF | .614 |
| CXCL12 | .586 |
| TNF-a | .748 |
| TNF-b | .668 |
| Extraction Method: Principal Component Analysis. | |
| a. 1 components extracted. | |

**Construction of PC06**

| **KMO and Bartlett's Test** | | | | |
| --- | --- | --- | --- | --- |
| Kaiser-Meyer-Olkin Measure of Sampling Adequacy. | | | | .866 |
| Bartlett's Test of Sphericity | | | Approx. Chi-Square | 794.053 |
|  |  |  | df | 45 |
|  |  |  | Sig. | .000 |
| **Component Matrix^a^** | |  |  |  |
|  | Component |  |  |  |
|  | 1 |  |  |  |
| sIL-1RA | .611 |  |  |  |
| IL-4 | .804 |  |  |  |
| IL-9 | .767 |  |  |  |
| IL-17 | .648 |  |  |  |
| CCL4 | .787 |  |  |  |
| PDGF | .749 |  |  |  |
| CCL5 | .777 |  |  |  |
| CXCL12 | .635 |  |  |  |
| TNF-a | .777 |  |  |  |
| TNF-b | .789 |  |  |  |
| Extraction Method: Principal Component Analysis. | |  |  |  |
| a. 1 components extracted. | |  |  |  |

**Construction of PCinv**

| **KMO and Bartlett's Test** | | |
| --- | --- | --- |
| Kaiser-Meyer-Olkin Measure of Sampling Adequacy. | | .715 |
| Bartlett's Test of Sphericity | Approx. Chi-Square | 220.576 |
|  | df | 28 |
|  | Sig. | .000 |

| **Component Matrix^a^** | |
| --- | --- |
|  | Component |
|  | 1 |
| IFN-a2 | .528 |
| IL-1a | .511 |
| IL-1b | .637 |
| IL-12p40 | .460 |
| IL-13 | .445 |
| NGF | .757 |
| VEGF | .778 |
| IL10dummy | .520 |
| Extraction Method: Principal Component Analysis. | |
| a. 1 components extracted. | |

**ESF, Table 5a. Differences in the four principal components between major depression (MDD, 1) and controls (noMDD, 0)**

| **Multivariate Tests^a^** | | | | | | |
| --- | --- | --- | --- | --- | --- | --- |
| Effect | | Value | F | Hypothesis df | Error df | Sig. |
| Intercept | Pillai's Trace | .015 | .481^b^ | 4.000 | 124.000 | .750 |
|  | Wilks' Lambda | .985 | .481^b^ | 4.000 | 124.000 | .750 |
|  | Hotelling's Trace | .016 | .481^b^ | 4.000 | 124.000 | .750 |
|  | Roy's Largest Root | .016 | .481^b^ | 4.000 | 124.000 | .750 |
| Sex | Pillai's Trace | .159 | 5.861^b^ | 4.000 | 124.000 | .000 |
|  | Wilks' Lambda | .841 | 5.861^b^ | 4.000 | 124.000 | .000 |
|  | Hotelling's Trace | .189 | 5.861^b^ | 4.000 | 124.000 | .000 |
|  | Roy's Largest Root | .189 | 5.861^b^ | 4.000 | 124.000 | .000 |
| Age | Pillai's Trace | .008 | .261^b^ | 4.000 | 124.000 | .902 |
|  | Wilks' Lambda | .992 | .261^b^ | 4.000 | 124.000 | .902 |
|  | Hotelling's Trace | .008 | .261^b^ | 4.000 | 124.000 | .902 |
|  | Roy's Largest Root | .008 | .261^b^ | 4.000 | 124.000 | .902 |
| MDD_NoMDD | Pillai's Trace | .286 | 12.431^b^ | 4.000 | 124.000 | .000 |
|  | Wilks' Lambda | .714 | 12.431^b^ | 4.000 | 124.000 | .000 |
|  | Hotelling's Trace | .401 | 12.431^b^ | 4.000 | 124.000 | .000 |
|  | Roy's Largest Root | .401 | 12.431^b^ | 4.000 | 124.000 | .000 |
| MetS_NoMetS | Pillai's Trace | .015 | .478^b^ | 4.000 | 124.000 | .752 |
|  | Wilks' Lambda | .985 | .478^b^ | 4.000 | 124.000 | .752 |
|  | Hotelling's Trace | .015 | .478^b^ | 4.000 | 124.000 | .752 |
|  | Roy's Largest Root | .015 | .478^b^ | 4.000 | 124.000 | .752 |
| MDD_NoMDD * MetS_NoMetS | Pillai's Trace | .158 | 5.801^b^ | 4.000 | 124.000 | .000 |
|  | Wilks' Lambda | .842 | 5.801^b^ | 4.000 | 124.000 | .000 |
|  | Hotelling's Trace | .187 | 5.801^b^ | 4.000 | 124.000 | .000 |
|  | Roy's Largest Root | .187 | 5.801^b^ | 4.000 | 124.000 | .000 |
| a. Design: Intercept + Sex + Age + MDD_NoMDD + MetS_NoMetS + MDD_NoMDD * MetS_NoMetS | | | | | | |
| b. Exact statistic | | | | | | |

**ESF, Table 5b. Measurements of the principal component scores in patients with major depression (MDD, 1) and controls (NoMDD, 0)**

| **Estimates** | | | | | |
| --- | --- | --- | --- | --- | --- |
| Dependent Variable | MDD_NoMDD | Mean | Std. Error | 95% Confidence Interval | |
|  |  |  |  | Lower Bound | Upper Bound |
| PC04 | .00 | -.164^a^ | .120 | -.401 | .072 |
|  | 1.00 | .178^a^ | .121 | -.061 | .417 |
| PC05 | .00 | -.306^a^ | .114 | -.531 | -.081 |
|  | 1.00 | .327^a^ | .115 | .099 | .554 |
| PC06 | .00 | -.311^a^ | .114 | -.536 | -.086 |
|  | 1.00 | .330^a^ | .115 | .103 | .558 |
| PCinv | .00 | .339^a^ | .113 | .115 | .563 |
|  | 1.00 | -.359^a^ | .114 | -.586 | -.133 |
| a. Covariates appearing in the model are evaluated at the following values: Sex = .203, Age = 37.436. | | | | | |

| **Univariate Tests** | | | | | | |
| --- | --- | --- | --- | --- | --- | --- |
| Dependent Variable | | Sum of Squares | df | Mean Square | F | Sig. |
| PC04 | Contrast | 3.760 | 1 | 3.760 | 3.982 | .048 |
|  | Error | 119.940 | 127 | .944 |  |  |
| PC05 | Contrast | 12.819 | 1 | 12.819 | 15.001 | .000 |
|  | Error | 108.526 | 127 | .855 |  |  |
| PC06 | Contrast | 13.173 | 1 | 13.173 | 15.427 | .000 |
|  | Error | 108.450 | 127 | .854 |  |  |
| PCinv | Contrast | 15.641 | 1 | 15.641 | 18.549 | .000 |
|  | Error | 107.090 | 127 | .843 |  |  |
| The F tests the effect of MDD_NoMDD. This test is based on the linearly independent pairwise comparisons among the estimated marginal means. | | | | | | |

**ESF, Table 5c. Differences in the four principal components between those with and without metabolic syndrome (MetS)**

| **Estimates** | | | | | |
| --- | --- | --- | --- | --- | --- |
| Dependent Variable | MetS_NoMetS | Mean | Std. Error | 95% Confidence Interval | |
|  |  |  |  | Lower Bound | Upper Bound |
| PC04 | .00 | .004^a^ | .125 | -.243 | .251 |
|  | 1.00 | .010^a^ | .130 | -.248 | .267 |
| PC05 | .00 | -.025^a^ | .119 | -.260 | .210 |
|  | 1.00 | .046^a^ | .124 | -.199 | .291 |
| PC06 | .00 | -.013^a^ | .119 | -.248 | .222 |
|  | 1.00 | .033^a^ | .124 | -.212 | .278 |
| PCinv | .00 | .072^a^ | .118 | -.161 | .306 |
|  | 1.00 | -.093^a^ | .123 | -.337 | .151 |
| a. Covariates appearing in the model are evaluated at the following values: Sex = .203, Age = 37.436. | | | | | |

| **Univariate Tests** | | | | | | |
| --- | --- | --- | --- | --- | --- | --- |
| Dependent Variable | | Sum of Squares | df | Mean Square | F | Sig. |
| PC04 | Contrast | .001 | 1 | .001 | .001 | .978 |
|  | Error | 119.940 | 127 | .944 |  |  |
| PC05 | Contrast | .131 | 1 | .131 | .154 | .696 |
|  | Error | 108.526 | 127 | .855 |  |  |
| PC06 | Contrast | .056 | 1 | .056 | .065 | .799 |
|  | Error | 108.450 | 127 | .854 |  |  |
| PCinv | Contrast | .702 | 1 | .702 | .833 | .363 |
|  | Error | 107.090 | 127 | .843 |  |  |

**ESF, Table 6a. Differences in cytokines between major depression (MDD, 1) and controls (noMDD, 0)**

| **Estimates** | | | | | |
| --- | --- | --- | --- | --- | --- |
| Dependent Variable | MDD_NoMDD | Mean | Std. Error | 95% Confidence Interval | |
|  |  |  |  | Lower Bound | Upper Bound |
| CCL27 | .00 | -.046^a^ | .119 | -.282 | .190 |
|  | 1.00 | .046^a^ | .122 | -.194 | .287 |
| CCL11 | .00 | -.268^a^ | .113 | -.492 | -.043 |
|  | 1.00 | .275^a^ | .116 | .046 | .504 |
| FGF | .00 | .164^a^ | .121 | -.076 | .404 |
|  | 1.00 | -.167^a^ | .124 | -.412 | .078 |
| G-CSF | .00 | .133^a^ | .121 | -.106 | .372 |
|  | 1.00 | -.138^a^ | .123 | -.382 | .106 |
| GM-CSF | .00 | .108^a^ | .121 | -.131 | .347 |
|  | 1.00 | -.149^a^ | .123 | -.393 | .095 |
| CXCL1 | .00 | -.098^a^ | .118 | -.332 | .137 |
|  | 1.00 | .051^a^ | .121 | -.188 | .290 |
| HGF | .00 | .023^a^ | .116 | -.207 | .253 |
|  | 1.00 | -.019^a^ | .119 | -.254 | .216 |
| IFN-a2 | .00 | .143^a^ | .118 | -.091 | .376 |
|  | 1.00 | -.150^a^ | .120 | -.388 | .089 |
| IFN-g | .00 | -.134^a^ | .121 | -.374 | .106 |
|  | 1.00 | .132^a^ | .124 | -.114 | .377 |
| IL-1a | .00 | .169^a^ | .121 | -.071 | .409 |
|  | 1.00 | -.189^a^ | .124 | -.435 | .056 |
| IL-1b | .00 | .219^a^ | .118 | -.015 | .452 |
|  | 1.00 | -.237^a^ | .120 | -.475 | .002 |
| sIL-1RA | .00 | -.178^a^ | .102 | -.380 | .025 |
|  | 1.00 | .177^a^ | .105 | -.030 | .384 |
| IL-2R | .00 | .010^a^ | .120 | -.227 | .248 |
|  | 1.00 | -.009^a^ | .123 | -.252 | .234 |
| IL-4 | .00 | -.260^a^ | .113 | -.484 | -.035 |
|  | 1.00 | .281^a^ | .116 | .052 | .510 |
| IL-8 | .00 | .162^a^ | .124 | -.083 | .407 |
|  | 1.00 | -.154^a^ | .126 | -.404 | .096 |
| IL-9 | .00 | -.299^a^ | .118 | -.532 | -.066 |
|  | 1.00 | .298^a^ | .120 | .060 | .536 |
| IL-12p40 | .00 | .205^a^ | .122 | -.036 | .447 |
|  | 1.00 | -.235^a^ | .125 | -.481 | .012 |
| IL-13 | .00 | .033^a^ | .120 | -.204 | .270 |
|  | 1.00 | -.057^a^ | .122 | -.299 | .185 |
| IL-16 | .00 | .043^a^ | .120 | -.194 | .280 |
|  | 1.00 | -.077^a^ | .122 | -.319 | .165 |
| IL-17 | .00 | -.148^a^ | .121 | -.387 | .091 |
|  | 1.00 | .151^a^ | .123 | -.093 | .395 |
| IL-18 | .00 | -.005^a^ | .123 | -.248 | .239 |
|  | 1.00 | .004^a^ | .126 | -.245 | .252 |
| CCXCL10 | .00 | -.092^a^ | .115 | -.319 | .135 |
|  | 1.00 | .059^a^ | .117 | -.173 | .291 |
| LIF | .00 | -.073^a^ | .121 | -.312 | .167 |
|  | 1.00 | .052^a^ | .123 | -.192 | .296 |
| CCL2 | .00 | -.208^a^ | .119 | -.443 | .027 |
|  | 1.00 | .217^a^ | .121 | -.023 | .457 |
| CCL7 | .00 | .139^a^ | .123 | -.105 | .383 |
|  | 1.00 | -.154^a^ | .126 | -.403 | .095 |
| M-CSF | .00 | .177^a^ | .118 | -.057 | .411 |
|  | 1.00 | -.182^a^ | .121 | -.420 | .057 |
| MIF | .00 | .113^a^ | .121 | -.128 | .353 |
|  | 1.00 | -.102^a^ | .124 | -.348 | .143 |
| CXCL9 | .00 | -.124^a^ | .122 | -.366 | .118 |
|  | 1.00 | .126^a^ | .125 | -.122 | .373 |
| CCL3 | .00 | .179^a^ | .120 | -.059 | .417 |
|  | 1.00 | -.176^a^ | .123 | -.419 | .067 |
| CCL4 | .00 | -.201^a^ | .120 | -.439 | .038 |
|  | 1.00 | .191^a^ | .123 | -.052 | .434 |
| NGF | .00 | .178^a^ | .120 | -.060 | .416 |
|  | 1.00 | -.196^a^ | .123 | -.439 | .046 |
| PDGF | .00 | -.285^a^ | .114 | -.511 | -.059 |
|  | 1.00 | .297^a^ | .117 | .067 | .528 |
| CCL5 | .00 | -.259^a^ | .108 | -.473 | -.046 |
|  | 1.00 | .242^a^ | .110 | .024 | .459 |
| SCF | .00 | .037^a^ | .114 | -.189 | .262 |
|  | 1.00 | -.021^a^ | .116 | -.251 | .209 |
| SCGF | .00 | -.063^a^ | .122 | -.304 | .177 |
|  | 1.00 | .073^a^ | .124 | -.172 | .319 |
| CXCL12 | .00 | -.353^a^ | .112 | -.575 | -.132 |
|  | 1.00 | .347^a^ | .114 | .121 | .573 |
| TNF-a | .00 | -.139^a^ | .122 | -.381 | .103 |
|  | 1.00 | .130^a^ | .125 | -.117 | .378 |
| TNF-B | .00 | -.408^a^ | .112 | -.630 | -.187 |
|  | 1.00 | .407^a^ | .114 | .181 | .633 |
| TRAIL | .00 | -.084^a^ | .117 | -.316 | .148 |
|  | 1.00 | .103^a^ | .119 | -.133 | .340 |
| VEGF | .00 | .276^a^ | .114 | .051 | .502 |
|  | 1.00 | -.291^a^ | .116 | -.522 | -.061 |
| a. Covariates appearing in the model are evaluated at the following values: Sex = .205, Age = 37.348, BMI = 27.1372. | | | | | |

| **Univariate Tests** | | | | | | |
| --- | --- | --- | --- | --- | --- | --- |
| Dependent Variable | | Sum of Squares | df | Mean Square | F | Sig. |
| CCL27 | Contrast | .269 | 1 | .269 | .288 | .593 |
|  | Error | 117.004 | 125 | .936 |  |  |
| CCL11 | Contrast | 9.331 | 1 | 9.331 | 11.038 | .001 |
|  | Error | 105.670 | 125 | .845 |  |  |
| FGF | Contrast | 3.472 | 1 | 3.472 | 3.588 | .060 |
|  | Error | 120.932 | 125 | .967 |  |  |
| G-CSF | Contrast | 2.332 | 1 | 2.332 | 2.426 | .122 |
|  | Error | 120.141 | 125 | .961 |  |  |
| GM-CSF | Contrast | 2.093 | 1 | 2.093 | 2.178 | .143 |
|  | Error | 120.093 | 125 | .961 |  |  |
| CXCL1 | Contrast | .699 | 1 | .699 | .757 | .386 |
|  | Error | 115.462 | 125 | .924 |  |  |
| HGF | Contrast | .056 | 1 | .056 | .063 | .802 |
|  | Error | 111.190 | 125 | .890 |  |  |
| IFN-a2 | Contrast | 2.709 | 1 | 2.709 | 2.954 | .088 |
|  | Error | 114.630 | 125 | .917 |  |  |
| IFN-g | Contrast | 2.239 | 1 | 2.239 | 2.311 | .131 |
|  | Error | 121.102 | 125 | .969 |  |  |
| IL-1a | Contrast | 4.067 | 1 | 4.067 | 4.190 | .043 |
|  | Error | 121.327 | 125 | .971 |  |  |
| IL-1b | Contrast | 6.573 | 1 | 6.573 | 7.170 | .008 |
|  | Error | 114.600 | 125 | .917 |  |  |
| sIL-1RA | Contrast | 3.977 | 1 | 3.977 | 5.760 | .018 |
|  | Error | 86.317 | 125 | .691 |  |  |
| IL-2R | Contrast | .012 | 1 | .012 | .012 | .912 |
|  | Error | 118.817 | 125 | .951 |  |  |
| IL-4 | Contrast | 9.249 | 1 | 9.249 | 10.940 | .001 |
|  | Error | 105.682 | 125 | .845 |  |  |
| IL-8 | Contrast | 3.159 | 1 | 3.159 | 3.135 | .079 |
|  | Error | 125.956 | 125 | 1.008 |  |  |
| IL-9 | Contrast | 11.282 | 1 | 11.282 | 12.348 | .001 |
|  | Error | 114.203 | 125 | .914 |  |  |
| IL-12p40 | Contrast | 6.125 | 1 | 6.125 | 6.248 | .014 |
|  | Error | 122.537 | 125 | .980 |  |  |
| IL-13 | Contrast | .260 | 1 | .260 | .275 | .601 |
|  | Error | 118.066 | 125 | .945 |  |  |
| IL-16 | Contrast | .455 | 1 | .455 | .482 | .489 |
|  | Error | 118.125 | 125 | .945 |  |  |
| IL-17 | Contrast | 2.824 | 1 | 2.824 | 2.946 | .089 |
|  | Error | 119.835 | 125 | .959 |  |  |
| IL-18 | Contrast | .002 | 1 | .002 | .002 | .962 |
|  | Error | 124.599 | 125 | .997 |  |  |
| CCXCL10 | Contrast | .718 | 1 | .718 | .829 | .364 |
|  | Error | 108.281 | 125 | .866 |  |  |
| LIF | Contrast | .492 | 1 | .492 | .511 | .476 |
|  | Error | 120.195 | 125 | .962 |  |  |
| CCL2 | Contrast | 5.719 | 1 | 5.719 | 6.165 | .014 |
|  | Error | 115.957 | 125 | .928 |  |  |
| CCL7 | Contrast | 2.723 | 1 | 2.723 | 2.726 | .101 |
|  | Error | 124.881 | 125 | .999 |  |  |
| M-CSF | Contrast | 4.065 | 1 | 4.065 | 4.418 | .038 |
|  | Error | 115.018 | 125 | .920 |  |  |
| MIF | Contrast | 1.465 | 1 | 1.465 | 1.509 | .222 |
|  | Error | 121.306 | 125 | .970 |  |  |
| CXCL9 | Contrast | 1.978 | 1 | 1.978 | 2.006 | .159 |
|  | Error | 123.276 | 125 | .986 |  |  |
| CCL3 | Contrast | 3.992 | 1 | 3.992 | 4.187 | .043 |
|  | Error | 119.184 | 125 | .953 |  |  |
| CCL4 | Contrast | 4.861 | 1 | 4.861 | 5.089 | .026 |
|  | Error | 119.392 | 125 | .955 |  |  |
| NGF | Contrast | 4.440 | 1 | 4.440 | 4.669 | .033 |
|  | Error | 118.860 | 125 | .951 |  |  |
| PDGF | Contrast | 10.728 | 1 | 10.728 | 12.488 | .001 |
|  | Error | 107.384 | 125 | .859 |  |  |
| CCL5 | Contrast | 7.947 | 1 | 7.947 | 10.399 | .002 |
|  | Error | 95.527 | 125 | .764 |  |  |
| SCF | Contrast | .106 | 1 | .106 | .125 | .725 |
|  | Error | 106.648 | 125 | .853 |  |  |
| SCGF | Contrast | .590 | 1 | .590 | .607 | .438 |
|  | Error | 121.534 | 125 | .972 |  |  |
| CXCL12 | Contrast | 15.555 | 1 | 15.555 | 18.892 | .000 |
|  | Error | 102.925 | 125 | .823 |  |  |
| TNF-a | Contrast | 2.300 | 1 | 2.300 | 2.331 | .129 |
|  | Error | 123.331 | 125 | .987 |  |  |
| TNF-B | Contrast | 21.050 | 1 | 21.050 | 25.500 | .000 |
|  | Error | 103.182 | 125 | .825 |  |  |
| TRAIL | Contrast | 1.111 | 1 | 1.111 | 1.232 | .269 |
|  | Error | 112.729 | 125 | .902 |  |  |
| VEGF | Contrast | 10.198 | 1 | 10.198 | 11.928 | .001 |
|  | Error | 106.872 | 125 | .855 |  |  |
| The F tests the effect of MDD_NoMDD. This test is based on the linearly independent pairwise comparisons among the estimated marginal means. | | | | | | |

See ESF, table 1 for explanation of the

**ESF, Table 6b. Differences in cytokines between those with (MetS, 1) and without (NoMetS, 0) metabolic syndrome**

| **Estimates** | | | | | |
| --- | --- | --- | --- | --- | --- |
| Dependent Variable | MetS_NoMetS | Mean | Std. Error | 95% Confidence Interval | |
|  |  |  |  | Lower Bound | Upper Bound |
| CCL27 | .00 | .123^a^ | .125 | -.124 | .369 |
|  | 1.00 | -.136^a^ | .130 | -.394 | .121 |
| CCL11 | .00 | .149^a^ | .118 | -.086 | .383 |
|  | 1.00 | -.158^a^ | .124 | -.403 | .086 |
| FGF | .00 | .043^a^ | .126 | -.206 | .292 |
|  | 1.00 | -.058^a^ | .131 | -.318 | .202 |
| G-CSF | .00 | .018^a^ | .129 | -.237 | .274 |
|  | 1.00 | -.022^a^ | .135 | -.288 | .245 |
| GM-CSF | .00 | .078^a^ | .126 | -.172 | .328 |
|  | 1.00 | -.093^a^ | .132 | -.353 | .168 |
| CXCL1 | .00 | -.090^a^ | .128 | -.343 | .162 |
|  | 1.00 | .096^a^ | .133 | -.168 | .360 |
| HGF | .00 | -.099^a^ | .126 | -.348 | .150 |
|  | 1.00 | .112^a^ | .132 | -.149 | .372 |
| IFN-a2 | .00 | .013^a^ | .124 | -.231 | .258 |
|  | 1.00 | -.029^a^ | .129 | -.284 | .226 |
| IFN-g | .00 | -.059^a^ | .127 | -.310 | .192 |
|  | 1.00 | .071^a^ | .132 | -.191 | .333 |
| IL-1a | .00 | .025^a^ | .127 | -.226 | .276 |
|  | 1.00 | -.030^a^ | .132 | -.292 | .231 |
| IL-1b | .00 | .051^a^ | .123 | -.192 | .293 |
|  | 1.00 | -.062^a^ | .128 | -.315 | .192 |
| sIL-1RA | .00 | -.147^a^ | .118 | -.382 | .087 |
|  | 1.00 | .174^a^ | .123 | -.070 | .418 |
| IL-2R | .00 | .008^a^ | .128 | -.246 | .262 |
|  | 1.00 | -.001^a^ | .134 | -.266 | .264 |
| IL-4 | .00 | .075^a^ | .118 | -.158 | .307 |
|  | 1.00 | -.062^a^ | .123 | -.305 | .181 |
| IL-8 | .00 | .003^a^ | .128 | -.251 | .257 |
|  | 1.00 | -.003^a^ | .134 | -.268 | .263 |
| IL-9 | .00 | -.080^a^ | .123 | -.323 | .163 |
|  | 1.00 | .094^a^ | .128 | -.160 | .348 |
| IL-12p40 | .00 | .028^a^ | .127 | -.223 | .279 |
|  | 1.00 | -.039^a^ | .132 | -.301 | .223 |
| IL-13 | .00 | .084^a^ | .124 | -.162 | .329 |
|  | 1.00 | -.099^a^ | .130 | -.355 | .157 |
| IL-16 | .00 | .025^a^ | .128 | -.229 | .278 |
|  | 1.00 | -.021^a^ | .134 | -.285 | .244 |
| IL-17 | .00 | .056^a^ | .125 | -.192 | .304 |
|  | 1.00 | -.049^a^ | .131 | -.307 | .210 |
| IL-18 | .00 | -.036^a^ | .130 | -.293 | .220 |
|  | 1.00 | .041^a^ | .135 | -.226 | .309 |
| CCXCL10 | .00 | -.116^a^ | .124 | -.361 | .129 |
|  | 1.00 | .132^a^ | .129 | -.124 | .388 |
| LIF | .00 | .062^a^ | .127 | -.190 | .313 |
|  | 1.00 | -.069^a^ | .133 | -.332 | .193 |
| CCL2 | .00 | -.004^a^ | .124 | -.249 | .241 |
|  | 1.00 | .019^a^ | .129 | -.237 | .275 |
| CCL7 | .00 | -.033^a^ | .128 | -.286 | .221 |
|  | 1.00 | .038^a^ | .134 | -.227 | .302 |
| M-CSF | .00 | .055^a^ | .124 | -.191 | .301 |
|  | 1.00 | -.052^a^ | .130 | -.308 | .205 |
| MIF | .00 | .037^a^ | .126 | -.212 | .286 |
|  | 1.00 | -.036^a^ | .131 | -.296 | .224 |
| CXCL9 | .00 | .019^a^ | .127 | -.232 | .270 |
|  | 1.00 | -.012^a^ | .132 | -.274 | .250 |
| CCL3 | .00 | -.014^a^ | .128 | -.267 | .240 |
|  | 1.00 | .010^a^ | .134 | -.254 | .275 |
| CCL4 | .00 | .019^a^ | .127 | -.232 | .269 |
|  | 1.00 | -.012^a^ | .132 | -.274 | .250 |
| NGF | .00 | -.002^a^ | .128 | -.255 | .250 |
|  | 1.00 | -.002^a^ | .133 | -.265 | .262 |
| PDGF | .00 | -.019^a^ | .119 | -.255 | .218 |
|  | 1.00 | .034^a^ | .125 | -.213 | .280 |
| CCL5 | .00 | -.017^a^ | .115 | -.244 | .210 |
|  | 1.00 | .040^a^ | .120 | -.197 | .277 |
| SCF | .00 | .129^a^ | .118 | -.106 | .363 |
|  | 1.00 | -.127^a^ | .124 | -.371 | .118 |
| SCGF | .00 | .020^a^ | .128 | -.234 | .273 |
|  | 1.00 | -.018^a^ | .134 | -.282 | .246 |
| CXCL12 | .00 | .049^a^ | .116 | -.181 | .280 |
|  | 1.00 | -.041^a^ | .122 | -.282 | .199 |
| TNF-a | .00 | .021^a^ | .127 | -.230 | .273 |
|  | 1.00 | -.013^a^ | .133 | -.275 | .250 |
| TNF-B | .00 | -.046^a^ | .117 | -.277 | .185 |
|  | 1.00 | .064^a^ | .122 | -.177 | .305 |
| TRAIL | .00 | -.054^a^ | .128 | -.306 | .199 |
|  | 1.00 | .064^a^ | .133 | -.200 | .328 |
| VEGF | .00 | .010^a^ | .120 | -.227 | .247 |
|  | 1.00 | -.027^a^ | .125 | -.275 | .220 |
| a. Covariates appearing in the model are evaluated at the following values: Sex = .203, Age = 37.436. | | | | | |

| **Univariate Tests** | | | | | | |
| --- | --- | --- | --- | --- | --- | --- |
| Dependent Variable | | Sum of Squares | df | Mean Square | F | Sig. |
| CCL27 | Contrast | 1.729 | 1 | 1.729 | 1.840 | .177 |
|  | Error | 119.331 | 127 | .940 |  |  |
| CCL11 | Contrast | 2.423 | 1 | 2.423 | 2.856 | .093 |
|  | Error | 107.729 | 127 | .848 |  |  |
| FGF | Contrast | .263 | 1 | .263 | .274 | .601 |
|  | Error | 122.015 | 127 | .961 |  |  |
| G-CSF | Contrast | .041 | 1 | .041 | .041 | .840 |
|  | Error | 128.001 | 127 | 1.008 |  |  |
| GM-CSF | Contrast | .747 | 1 | .747 | .775 | .380 |
|  | Error | 122.500 | 127 | .965 |  |  |
| CXCL1 | Contrast | .891 | 1 | .891 | .903 | .344 |
|  | Error | 125.286 | 127 | .987 |  |  |
| HGF | Contrast | 1.140 | 1 | 1.140 | 1.186 | .278 |
|  | Error | 122.075 | 127 | .961 |  |  |
| IFN-a2 | Contrast | .046 | 1 | .046 | .050 | .823 |
|  | Error | 117.309 | 127 | .924 |  |  |
| IFN-g | Contrast | .433 | 1 | .433 | .445 | .506 |
|  | Error | 123.577 | 127 | .973 |  |  |
| IL-1a | Contrast | .079 | 1 | .079 | .081 | .776 |
|  | Error | 123.434 | 127 | .972 |  |  |
| IL-1b | Contrast | .324 | 1 | .324 | .356 | .552 |
|  | Error | 115.541 | 127 | .910 |  |  |
| sIL-1RA | Contrast | 2.656 | 1 | 2.656 | 3.134 | .079 |
|  | Error | 107.620 | 127 | .847 |  |  |
| IL-2R | Contrast | .002 | 1 | .002 | .002 | .961 |
|  | Error | 126.669 | 127 | .997 |  |  |
| IL-4 | Contrast | .479 | 1 | .479 | .572 | .451 |
|  | Error | 106.210 | 127 | .836 |  |  |
| IL-8 | Contrast | .001 | 1 | .001 | .001 | .978 |
|  | Error | 126.735 | 127 | .998 |  |  |
| IL-9 | Contrast | .774 | 1 | .774 | .847 | .359 |
|  | Error | 116.052 | 127 | .914 |  |  |
| IL-12p40 | Contrast | .118 | 1 | .118 | .121 | .729 |
|  | Error | 123.722 | 127 | .974 |  |  |
| IL-13 | Contrast | .858 | 1 | .858 | .920 | .339 |
|  | Error | 118.404 | 127 | .932 |  |  |
| IL-16 | Contrast | .053 | 1 | .053 | .053 | .818 |
|  | Error | 126.084 | 127 | .993 |  |  |
| IL-17 | Contrast | .281 | 1 | .281 | .296 | .587 |
|  | Error | 120.836 | 127 | .951 |  |  |
| IL-18 | Contrast | .155 | 1 | .155 | .152 | .697 |
|  | Error | 129.119 | 127 | 1.017 |  |  |
| CCXCL10 | Contrast | 1.578 | 1 | 1.578 | 1.697 | .195 |
|  | Error | 118.094 | 127 | .930 |  |  |
| LIF | Contrast | .443 | 1 | .443 | .453 | .502 |
|  | Error | 124.171 | 127 | .978 |  |  |
| CCL2 | Contrast | .014 | 1 | .014 | .015 | .902 |
|  | Error | 117.960 | 127 | .929 |  |  |
| CCL7 | Contrast | .128 | 1 | .128 | .129 | .721 |
|  | Error | 126.264 | 127 | .994 |  |  |
| M-CSF | Contrast | .293 | 1 | .293 | .313 | .577 |
|  | Error | 118.765 | 127 | .935 |  |  |
| MIF | Contrast | .139 | 1 | .139 | .145 | .704 |
|  | Error | 121.759 | 127 | .959 |  |  |
| CXCL9 | Contrast | .026 | 1 | .026 | .026 | .871 |
|  | Error | 123.736 | 127 | .974 |  |  |
| CCL3 | Contrast | .015 | 1 | .015 | .015 | .902 |
|  | Error | 125.989 | 127 | .992 |  |  |
| CCL4 | Contrast | .024 | 1 | .024 | .025 | .874 |
|  | Error | 123.443 | 127 | .972 |  |  |
| NGF | Contrast | 2.141E-5 | 1 | 2.141E-5 | .000 | .996 |
|  | Error | 125.531 | 127 | .988 |  |  |
| PDGF | Contrast | .071 | 1 | .071 | .082 | .775 |
|  | Error | 109.629 | 127 | .863 |  |  |
| CCL5 | Contrast | .084 | 1 | .084 | .105 | .747 |
|  | Error | 101.523 | 127 | .799 |  |  |
| SCF | Contrast | 1.675 | 1 | 1.675 | 1.973 | .163 |
|  | Error | 107.830 | 127 | .849 |  |  |
| SCGF | Contrast | .036 | 1 | .036 | .036 | .849 |
|  | Error | 125.925 | 127 | .992 |  |  |
| CXCL12 | Contrast | .212 | 1 | .212 | .258 | .612 |
|  | Error | 104.341 | 127 | .822 |  |  |
| TNF-a | Contrast | .030 | 1 | .030 | .031 | .861 |
|  | Error | 124.358 | 127 | .979 |  |  |
| TNF-B | Contrast | .313 | 1 | .313 | .379 | .539 |
|  | Error | 104.807 | 127 | .825 |  |  |
| TRAIL | Contrast | .353 | 1 | .353 | .357 | .551 |
|  | Error | 125.594 | 127 | .989 |  |  |
| VEGF | Contrast | .037 | 1 | .037 | .042 | .838 |
|  | Error | 110.433 | 127 | .870 |  |  |
| The F tests the effect of MetS_NoMetS. This test is based on the linearly independent pairwise comparisons among the estimated marginal means. | | | | | | |

**ESF, Table 6c. Associations between body mass index (BMI) and cytokines**

|  | Dependent Variable | Sum of Squares | df | Mean Square | F | Sig. |
| --- | --- | --- | --- | --- | --- | --- |
| BMI | CCL27 | 2.173 | 1 | 2.173 | 2.322 | .130 |
|  | CCL11 | 1.382 | 1 | 1.382 | 1.635 | .203 |
|  | FGF | .196 | 1 | .196 | .202 | .654 |
|  | G-CSF | 7.080 | 1 | 7.080 | 7.366 | .008 |
|  | GM-CSF | .019 | 1 | .019 | .020 | .887 |
|  | CXCL1 | .373 | 1 | .373 | .404 | .526 |
|  | HGF | 10.495 | 1 | 10.495 | 11.798 | .001 |
|  | IFN-a2 | 2.677 | 1 | 2.677 | 2.919 | .090 |
|  | IFN-g | 2.197 | 1 | 2.197 | 2.268 | .135 |
|  | IL-1a | 1.760 | 1 | 1.760 | 1.813 | .181 |
|  | IL-1b | .302 | 1 | .302 | .330 | .567 |
|  | sIL-1RA | 21.289 | 1 | 21.289 | 30.830 | .000 |
|  | IL-2R | 7.481 | 1 | 7.481 | 7.870 | .006 |
|  | IL-4 | .049 | 1 | .049 | .058 | .811 |
|  | IL-8 | .386 | 1 | .386 | .383 | .537 |
|  | IL-9 | 1.620 | 1 | 1.620 | 1.773 | .185 |
|  | IL-12p40 | .081 | 1 | .081 | .083 | .774 |
|  | IL-13 | .155 | 1 | .155 | .164 | .686 |
|  | IL-16 | 5.569 | 1 | 5.569 | 5.893 | .017 |
|  | IL-17 | .946 | 1 | .946 | .987 | .323 |
|  | IL-18 | 4.441 | 1 | 4.441 | 4.455 | .037 |
|  | CXCL10 | 5.253 | 1 | 5.253 | 6.064 | .015 |
|  | LIF | 3.961 | 1 | 3.961 | 4.119 | .045 |
|  | CCL2 | 1.980 | 1 | 1.980 | 2.134 | .147 |
|  | CCL7 | .122 | 1 | .122 | .123 | .727 |
|  | M-CSF | 3.704 | 1 | 3.704 | 4.025 | .047 |
|  | MIF | .119 | 1 | .119 | .123 | .727 |
|  | CXCL9 | .451 | 1 | .451 | .458 | .500 |
|  | CCL3 | 5.368 | 1 | 5.368 | 5.630 | .019 |
|  | CCL4 | 3.979 | 1 | 3.979 | 4.166 | .043 |
|  | NGF | 3.723 | 1 | 3.723 | 3.915 | .050 |
|  | PDGF | 2.192 | 1 | 2.192 | 2.552 | .113 |
|  | CCL5 | .991 | 1 | .991 | 1.297 | .257 |
|  | SCF | .978 | 1 | .978 | 1.147 | .286 |
|  | SCGF | 3.245 | 1 | 3.245 | 3.337 | .070 |
|  | CXCL12 | .228 | 1 | .228 | .277 | .599 |
|  | TNF-a | .530 | 1 | .530 | .537 | .465 |
|  | TNF-B | 1.007 | 1 | 1.007 | 1.220 | .272 |
|  | TRAIL | 10.394 | 1 | 10.394 | 11.526 | .001 |
|  | VEGF | 3.099 | 1 | 3.099 | 3.624 | .059 |

**ESF, Table 7 Interaction patterns between major depression (MDD) and metabolic syndrome (MetS).**

|  |  | Type III sum of squares | df | Mean square | F | p |
| --- | --- | --- | --- | --- | --- | --- |
| MDD* MetS_ | IL-1signaling | 3.719 | 1/127 | 3.719 | 4.590 | .034 |
|  | TNF signaling | 5.034 | 1/127 | 5.034 | 5.494 | .021 |
|  | PC04 | 7.723 | 1/127 | 7.723 | 8.178 | .005 |
|  | PC05 | 11.480 | 1/127 | 11.480 | 13.435 | .000 |
|  | PC06 | 10.976 | 1/127 | 10.976 | 12.854 | .000 |
|  | PCinv | 5.198 | 1/127 | 5.198 | 6.164 | .014 |

See table 2 for explanation of interkeukin-1 (IL_ and tumor necrosis factor (TNF) signaling.

**ESF, Figure 1. Effects of interaction between major depression (MDD) X metabolic syndrome (MetS) on interleukin-1 signaling.** See ESF, Tables 2 and 4 for explanation. See ESF, Table 7 for statistics.

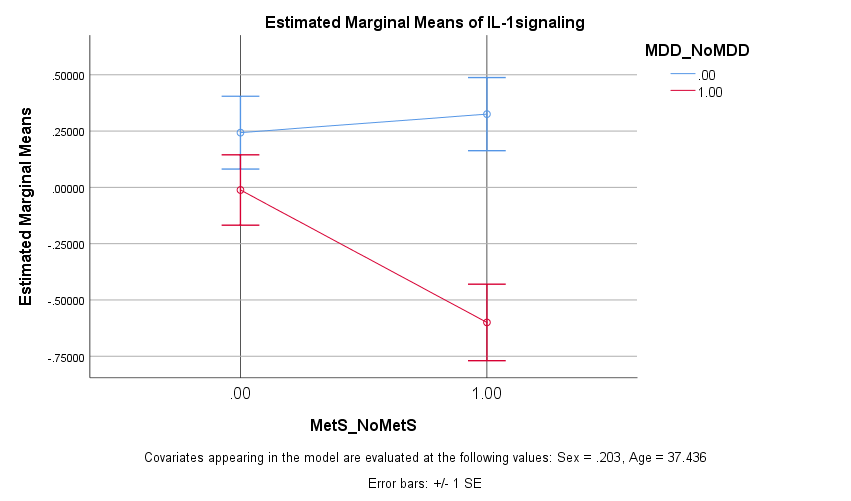

**ESF, Figure 2. Effects of interaction major depression (MDD) X metabolic syndrome (MetS) on tumor necrosis factor (TNF) signaling.** See ESF, Tables 2 and 4 for explanation. See ESF, Table 7 for statistics.

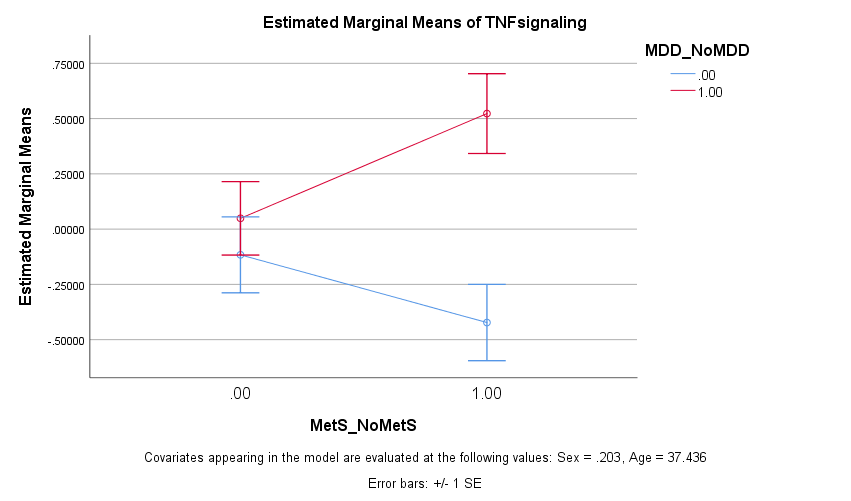

**ESF, Figure 3. Effects of interaction major depression (MDD) X metabolic syndrome (MetS) on principal component 04 (PC04).** See ESF, Tables 4 for explanation. See ESF, Table 7 for statistics.

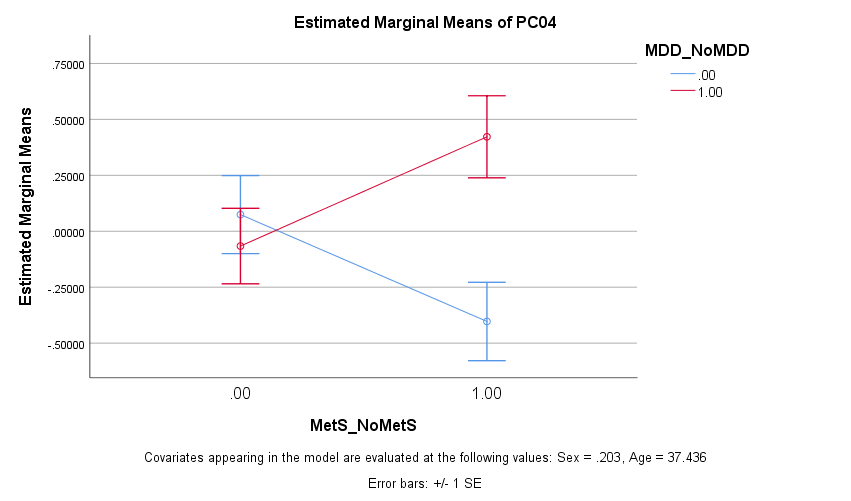

**ESF, Figure 4. Effects of interaction major depression (MDD) X metabolic syndrome (MetS) on principal component 05 (PC05).** See ESF, Tables 4 for explanation. See ESF, Table 7 for statistics.

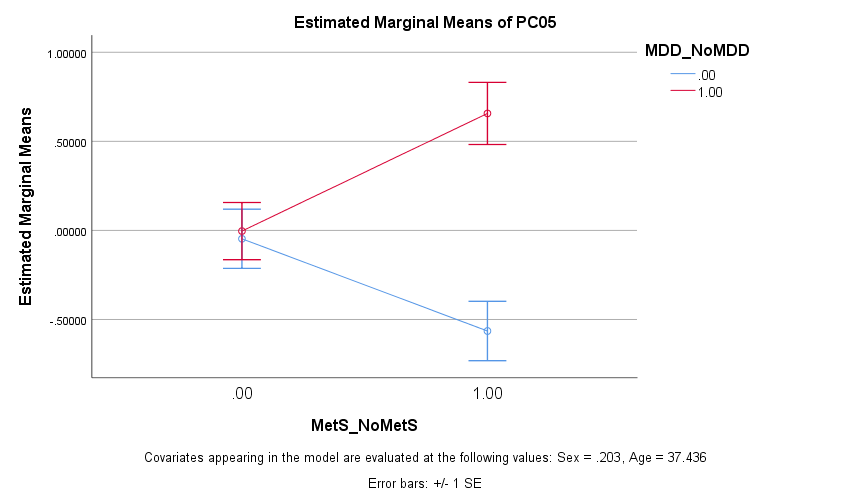

**ESF, Figure 5. Effects of interaction major depression (MDD) X metabolic syndrome (MetS) on principal component 05 (PC05).** See ESF, Tables 4 for explanation. See ESF, Table 7 for statistics.

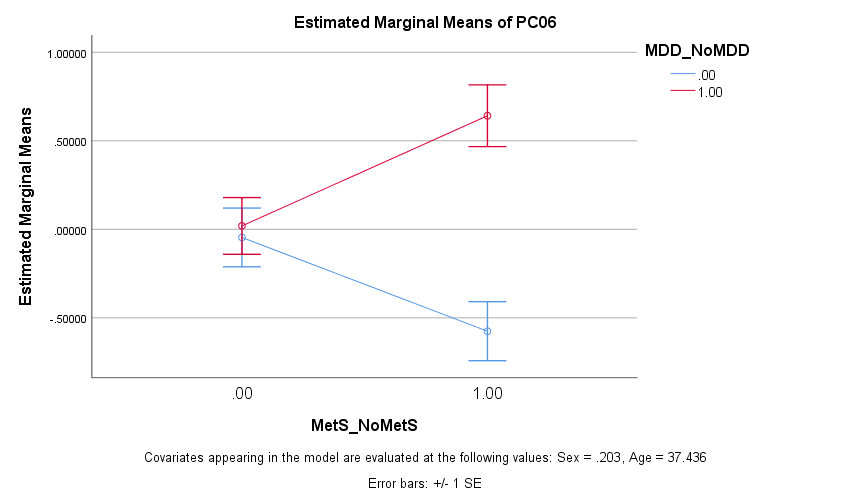

**ESF, Figure 6. Effects of interaction major depression (MDD) X metabolic syndrome (MetS) on principal component inv (PCinv).** See ESF, Tables 4 for explanation. See ESF, Table 7 for statistics.

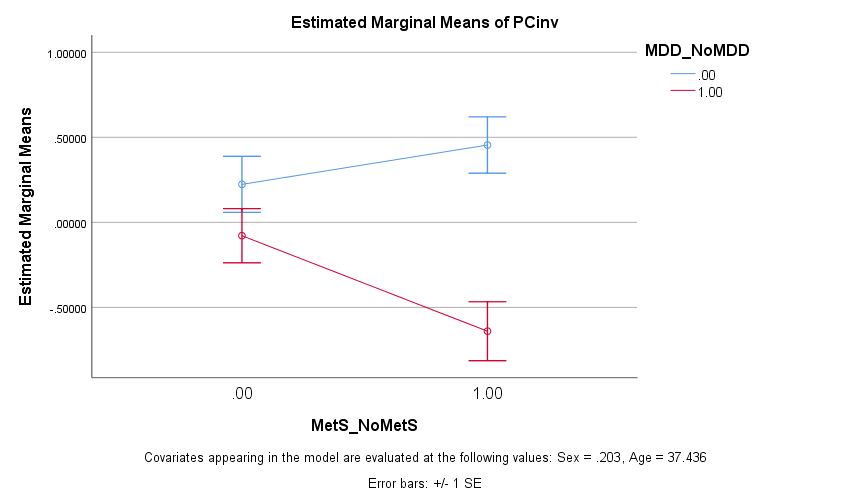

**ESF, Table 8. Interaction patterns between major depression (MDD) and metabolic syndrome (MetS) on serum cytokine levels.**

| Source | Dependent Variable | Type III Sum of Squares | df | Mean Square | F | Sig. |
| --- | --- | --- | --- | --- | --- | --- |
| MDD_* MetS | FGF | 4.725 | 1 | 4.725 | 4.918 | .028 |
|  | IFN-a2 | 9.226 | 1 | 9.226 | 9.989 | .002 |
|  | sIL-1RA | 8.896 | 1 | 8.896 | 10.498 | .002 |
|  | IL-2R | 3.960 | 1 | 3.960 | 3.970 | .048 |
|  | IL-4 | 12.745 | 1 | 12.745 | 15.240 | .000 |
|  | IL-13 | 4.186 | 1 | 4.186 | 4.490 | .036 |
|  | IL-17 | 5.608 | 1 | 5.608 | 5.894 | .017 |
|  | CCL2 | 7.409 | 1 | 7.409 | 7.977 | .006 |
|  | M-CSF | 6.567 | 1 | 6.567 | 7.022 | .009 |
|  | PDGF | 4.823 | 1 | 4.823 | 5.587 | .020 |
|  | CCL5 | 18.450 | 1 | 18.450 | 23.080 | .000 |
|  | SCF | 9.362 | 1 | 9.362 | 11.026 | .001 |
|  | TNF-a | 3.900 | 1 | 3.900 | 3.983 | .048 |
|  | TNF-B | 4.086 | 1 | 4.086 | 4.951 | .028 |
|  | VEGF | 8.855 | 1 | 8.855 | 10.184 | .002 |

See Table 1 for explanations

**ESF, Figure 7. Effects of interaction major depression (MDD) X metabolic syndrome (MetS) on fibroblast growth factor (FGF).** See ESF, Table 7 for statistics.

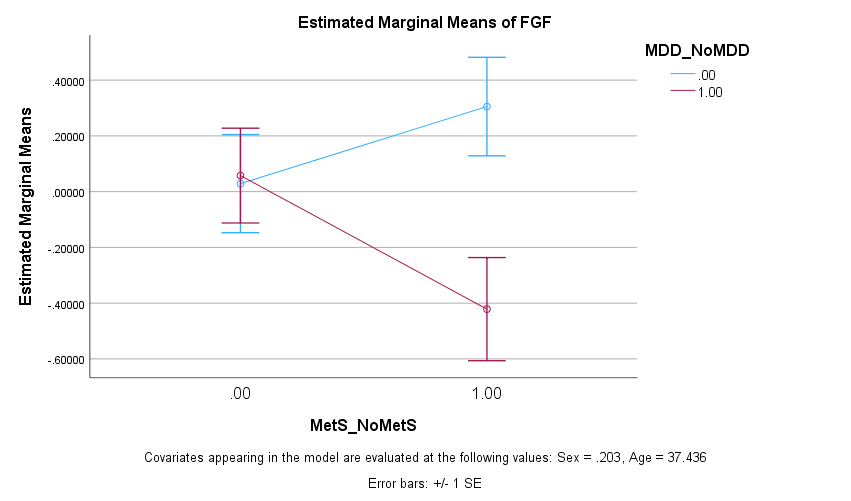

**ESF, Figure 8. Effects of interaction major depression (MDD) X metabolic syndrome (MetS) on interferon alpha-2 (IFN-a2).** See ESF, Table 7 for statistics.

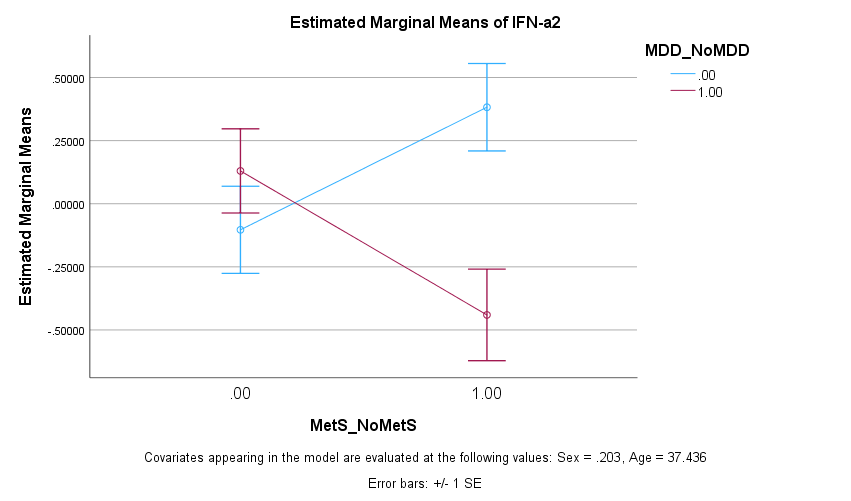

**ESF, Figure 9. Effects of interaction major depression (MDD) X metabolic syndrome (MetS) on soluble interleukin-1 receptor antagonist (sIL-1RA).** See ESF, Table 7 for statistics.

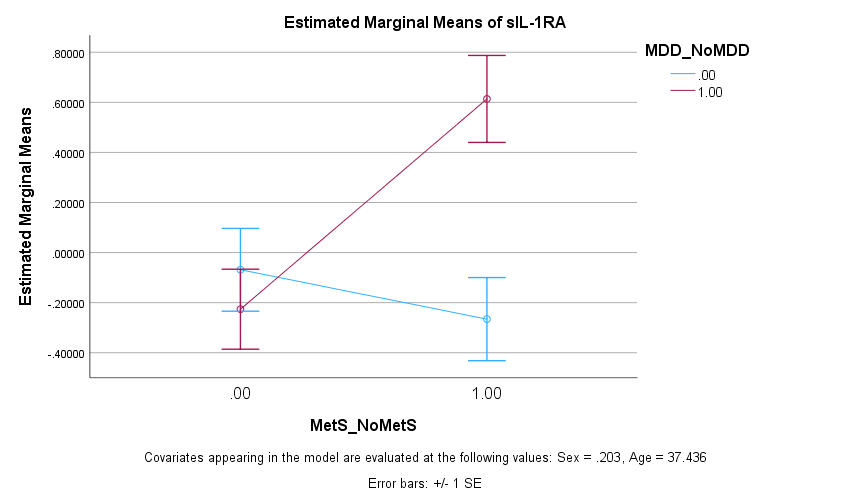

**ESF, Figure 10. Effects of interaction major depression (MDD) X metabolic syndrome (MetS) on soluble interleukin-2 receptor (sIL-2R).** See ESF, Table 7 for statistics.

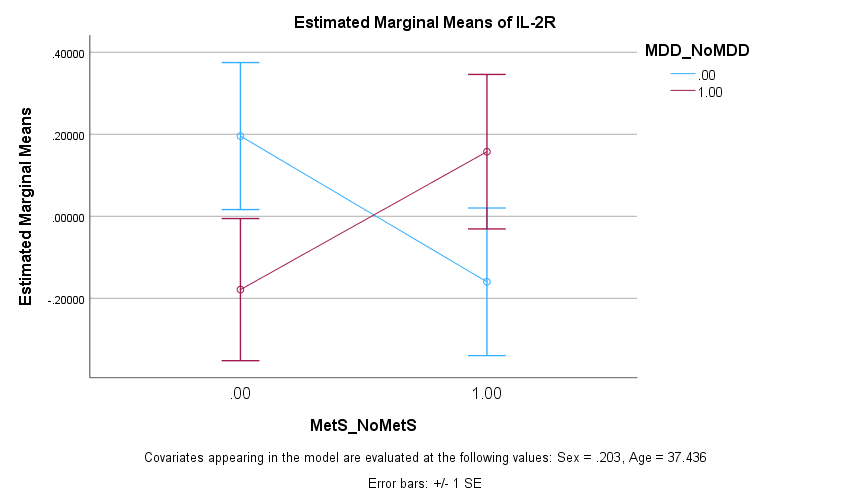

**ESF, Figure 11. Effects of interaction major depression (MDD) X metabolic syndrome (MetS) on soluble interleukin-4 (IL-4).** See ESF, Table 7 for statistics.

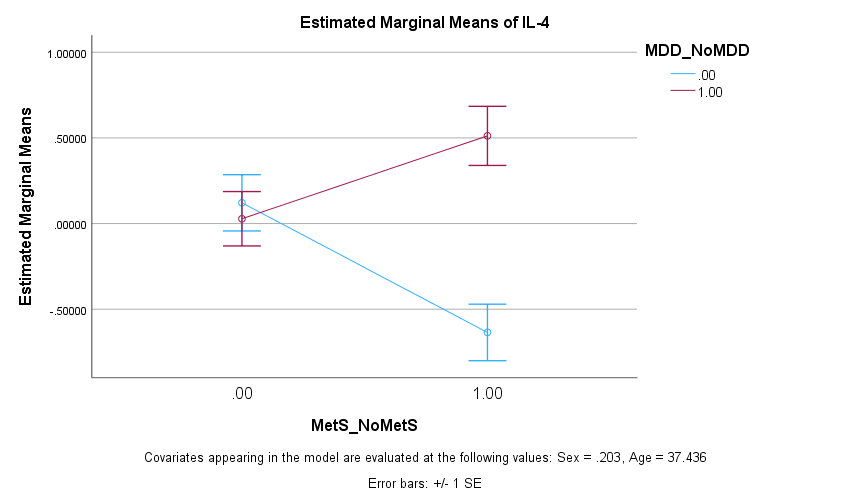

**ESF, Figure 12. Effects of interaction major depression (MDD) X metabolic syndrome (MetS) on soluble interleukin-13 (IL-13).** See ESF, Table 7 for statistics.

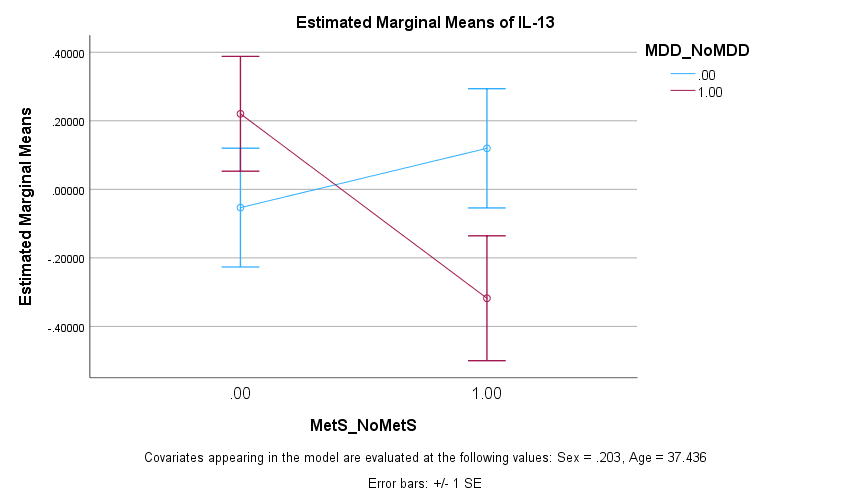

**ESF, Figure 13. Effects of interaction major depression (MDD) X metabolic syndrome (MetS) on soluble interleukin-13 (IL-13).** See ESF, Table 7 for statistics.

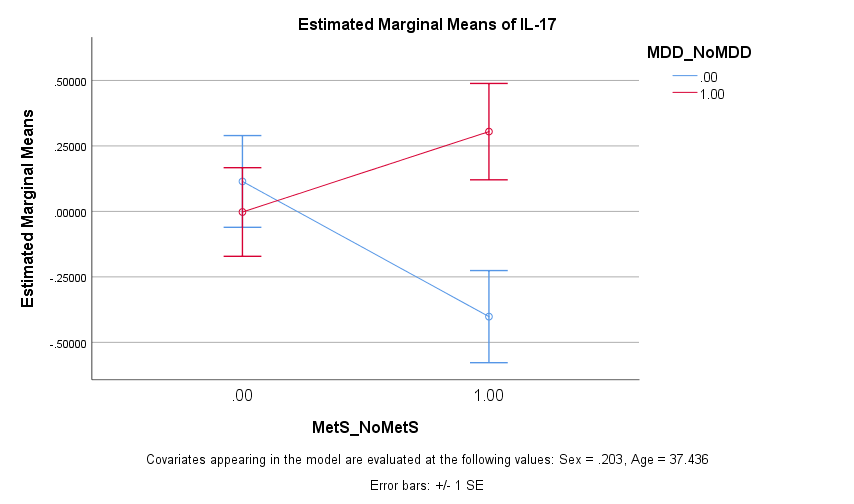

**ESF, Figure 14. Effects of interaction major depression (MDD) X metabolic syndrome (MetS) on CCL2.** See ESF, Table 7 for statistics.

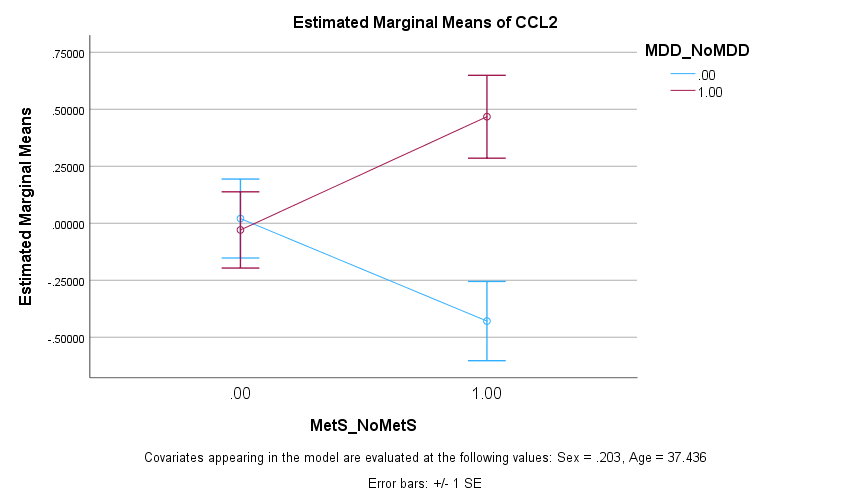

**ESF, Figure 15. Effects of interaction major depression (MDD) X metabolic syndrome (MetS) on macrophage-colony stimulating factor (M-CSF).** See ESF, Table 7 for statistics.

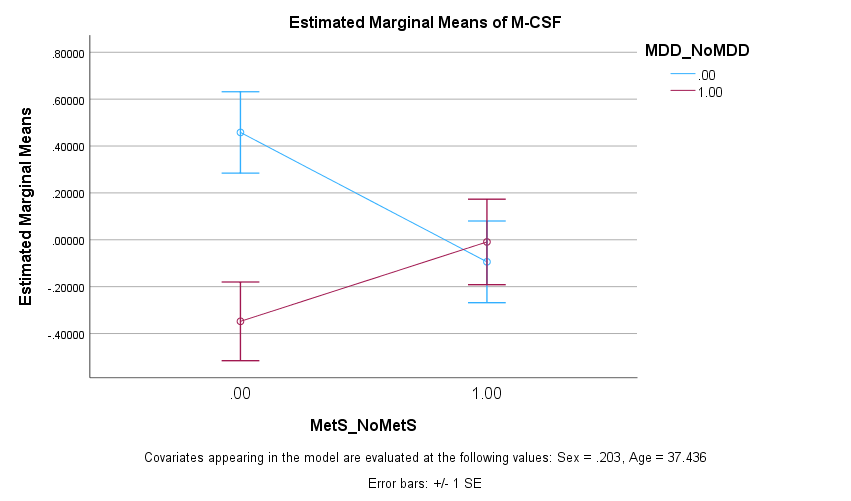

**ESF, Figure 16. Effects of interaction major depression (MDD) X metabolic syndrome (MetS) on platelet-derived growth factor (PDGF).** See ESF, Table 7 for statistics.

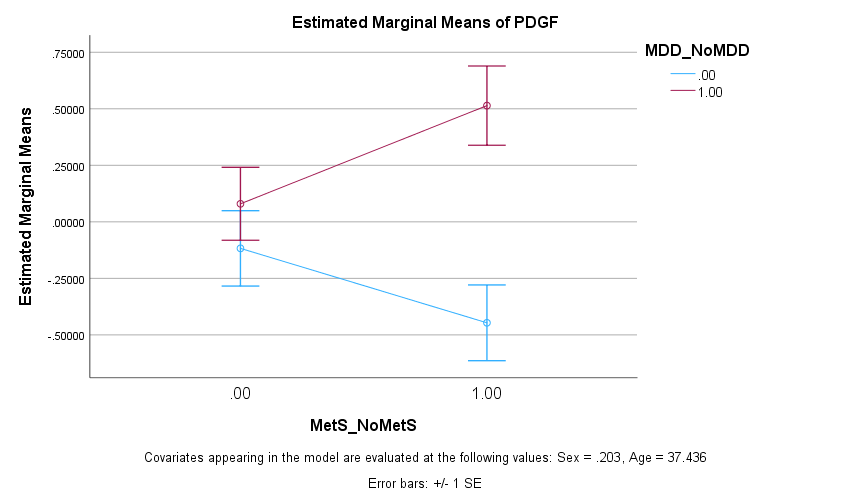

**ESF, Figure 17. Effects of interaction major depression (MDD) X metabolic syndrome (MetS) on CCL5 (CCL5).** See ESF, Table 7 for statistics.

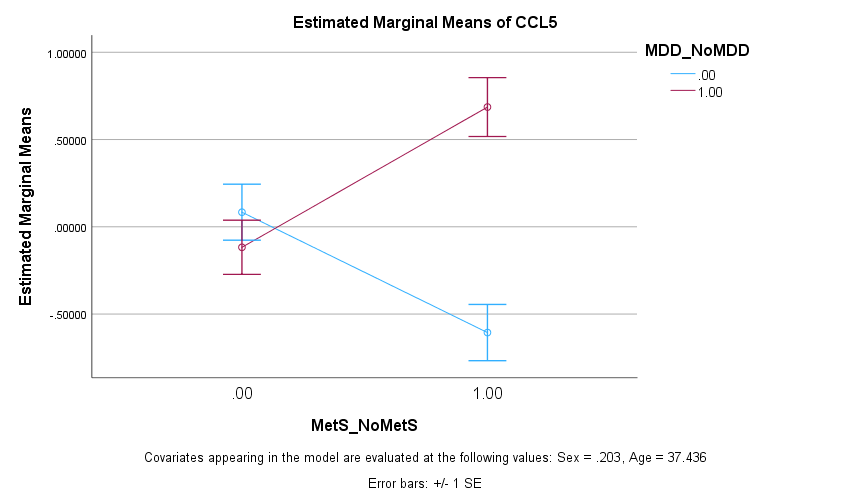

**ESF, Figure 18. Effects of interaction major depression (MDD) X metabolic syndrome (MetS) on stem cell factor (SCF).** See ESF, Table 7 for statistics.

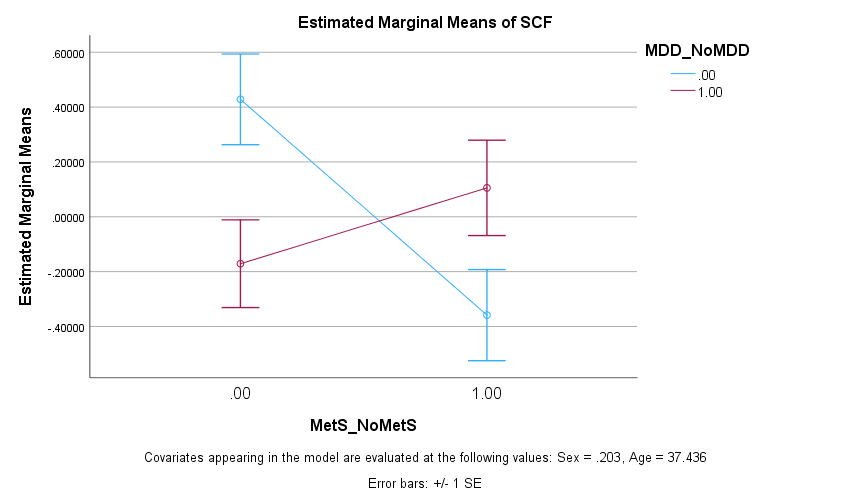

**ESF, Figure 19. Effects of interaction major depression (MDD) X metabolic syndrome (MetS) on tumor necrosis factor alpha (TNF-a).** See ESF, Table 7 for statistics.

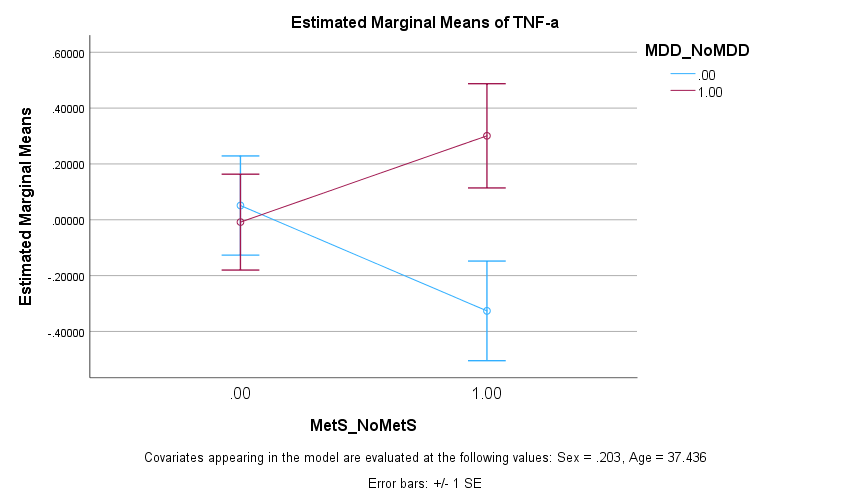

**ESF, Figure 20. Effects of interaction major depression (MDD) X metabolic syndrome (MetS) on tumor necrosis factor beta (TNF-β).** See ESF, Table 7 for statistics.

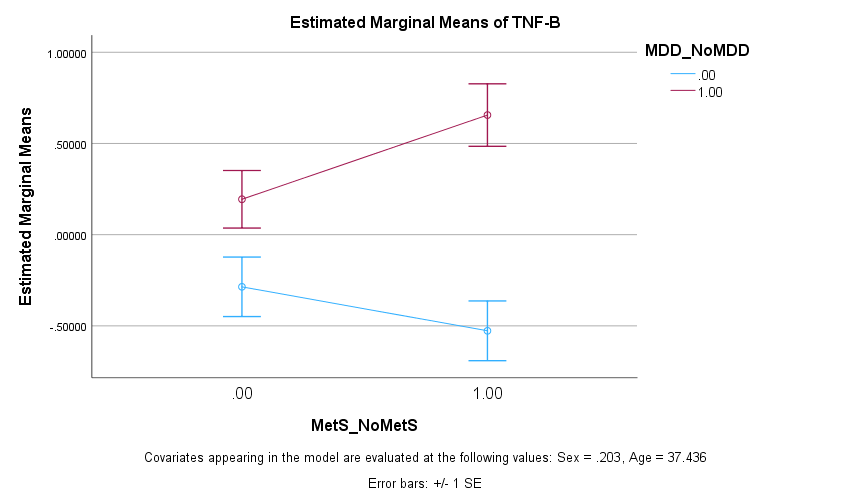

**ESF, Figure 21. Effects of interaction major depression (MDD) X metabolic syndrome (MetS) on vascular endothelial growth factor (VEGF).** See ESF, Table 7 for statistics.

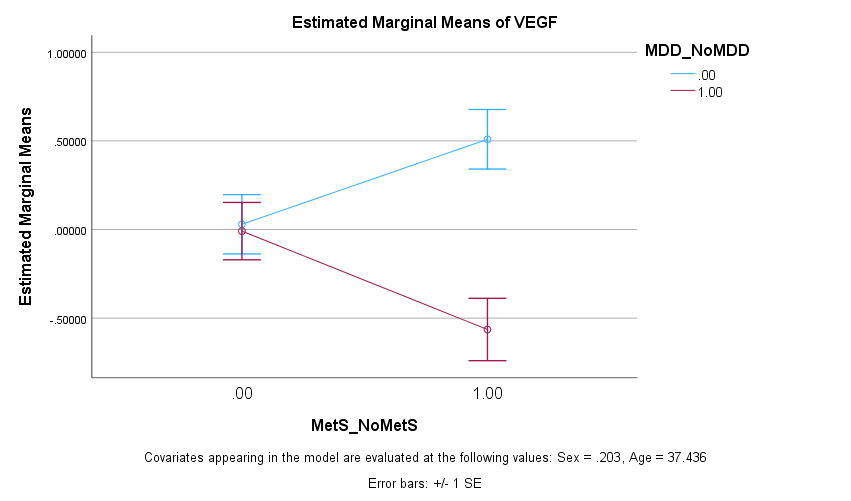

**ESF, Table 9. Biological GO term classifications of upregulated differently expressed proteins in major depression.**

| **ID term** | **Term description** | **Observed** | **Background** | **Strength** | **p FDR** |
| --- | --- | --- | --- | --- | --- |
| GO:0006955 | Immune response | 17 | 1321 | 1.1 | 1.83e-13 |
| GO:0006954 | Inflammatory response | 13 | 538 | 1.38 | 1.96e-12 |
| GO:0019221 | Cytokine-mediated signaling pathway | 12 | 369 | 1.51 | 1.96e-12 |
| GO:0032101 | Regulation of response to external stimulus | 15 | 964 | 1.19 | 1.96e-12 |
| GO:0050900 | Leukocyte migration | 10 | 249 | 1.6 | 6.63e-11 |
| GO:0006952 | Defense response | 15 | 1394 | 1.03 | 1.49e-10 |
| GO:0002684 | Positive regulation of immune system process | 13 | 874 | 1.17 | 2.96e-10 |
| GO:0032103 | Positive regulation of response to external stimulus | 11 | 453 | 1.38 | 2.96e-10 |
| GO:0060326 | Cell chemotaxis | 9 | 210 | 1.63 | 5.17e-10 |
| GO:0007165 | Signal transduction | 20 | 4714 | 0.62 | 5.59e-10 |
| GO:0002688 | Regulation of leukocyte chemotaxis | 8 | 128 | 1.79 | 6.38e-10 |
| GO:0002685 | Regulation of leukocyte migration | 9 | 231 | 1.58 | 6.67e-10 |

See ESF, Table 4 for the cytokines/chemokines/growth factors that are upregulated in principal component PC05.

Results of Gene Ontology (GO) biological process functional enrichment analysis (using STRING).

p FDR: False discovery rate p correction.

**ESF, Table 10.** **Biological GO term classifications of upregulated differently expressed proteins in major depression.**

| **ID term** | **Term description** | **Observed** | **Background** | **Strength** | **p FDR** |
| --- | --- | --- | --- | --- | --- |
| GO:0048247 | Lymphocyte chemotaxis | 5 | 51 | 2.29 | 6.03e-07 |
| GO:0006952 | Defense response | 9 | 1394 | 1.1 | 1.56e-06 |
| GO:0006955 | Immune response | 9 | 1321 | 1.13 | 1.56e-06 |
| GO:0048245 | Eosinophil chemotaxis | 4 | 17 | 2.67 | 1.56e-06 |
| GO:0070098 | Chemokine-mediated signaling pathway | 5 | 82 | 2.08 | 1.56e-06 |
| GO:0006954 | Inflammatory response | 7 | 538 | 1.41 | 2.08e-06 |
| GO:0071675 | Regulation of mononuclear cell migration | 5 | 125 | 1.9 | 3.71e-06 |
| GO:0002688 | Regulation of leukocyte chemotaxis | 5 | 128 | 1.89 | 3.84e-06 |
| GO:0002548 | Monocyte chemotaxis | 4 | 43 | 2.26 | 6.61e-06 |
| GO:2000501 | Regulation of natural killer cell chemotaxis | 3 | 8 | 2.87 | 1.52e-05 |
| GO:0006950 | Response to stress | 10 | 3358 | 0.77 | 1.83e-05 |
| GO:0002676 | Regulation of chronic inflammatory response | 3 | 10 | 2.77 | 2.21e-05 |

See Figure 1 for the cytokines/chemokines/growth factors that are upregulated in major depression (CXCL12, TNFB, PDGF, CCL11, IL9, IL4, CCL5, CCL2, CCL4, and IL1RA).

Results of Gene Ontology (GO) biological process functional enrichment analysis (using STRING).

p FDR: False discovery rate p correction.

**ESF, Table 11. Important biological GO term classifications of downregulated differently expressed proteins in major depression.**

| **ID term** | **Term description** | **p** | **p FDR** | **Number of DEPs** | **Genes** |
| --- | --- | --- | --- | --- | --- |
| GO:0009967 | positive regulation of signal transduction | 2.72E-08 | 7.03E-05 | 7 | CCL3\|CSF1\|IL10\|IL12B\|IL1B\|NGF\|VEGFA |
| GO:0010941 | regulation of cell death | 3.07E-08 | 7.03E-05 | 7 | CCL3\|CSF1\|IL10\|IL12B\|IL1B\|NGF\|VEGFA |
| GO:0010647 | positive regulation of cell communication | 4.95E-08 | 7.03E-05 | 7 | CCL3\|CSF1\|IL10\|IL12B\|IL1B\|NGF\|VEGFA |
| GO:0045785 | positive regulation of cell adhesion | 6.54E-08 | 8.01E-05 | 5 | CSF1\|IL10\|IL12B\|IL1B\|VEGFA |
| GO:0045597 | positive regulation of cell differentiation | 8.18E-08 | 8.64E-05 | 6 | CCL3\|CSF1\|IL12B\|IL1B\|NGF\|VEGFA |
| GO:0050769 | positive regulation of neurogenesis | 1.38E-07 | 9.63E-05 | 5 | CCL3\|CSF1\|IL1B\|NGF\|VEGFA |
| GO:0022603 | regulation of anatomical structure morphogenesis | 2.22E-08 | 2.09E-04 | 6 | CCL3\|CSF1\|IL10\|IL1B\|NGF\|VEGFA |
| GO:0051962 | positive regulation of nervous system development | 7.890E-08 | 2.092E-04 | 5 | CCL3\|CSF1\|IL1B\|NGF\|VEGFA |
| GO:0010720 | positive regulation of cell development | 8.190E-08 | 2.092E-04 | 5 | CCL3\|CSF1\|IL1B\|NGF\|VEGFA |
| GO:0060326 | cell chemotaxis | 1.755E-07 | 3.223E-04 | 4 | CCL3\|IL10\|IL1B\|VEGFA |
| GO:0019221 | cytokine-mediated signaling pathway | 2.435E-07 | 3.834E-04 | 5 | CCL3\|CSF1\|IL10\|IL1B\|VEGFA |

See Figure 1 for the downregulated differentially expressed proteins (NGF, IL1B, CSF1, CCL3, IL12B, VEGFA, IL10).

Results of Gene Ontology (GO) biological process functional enrichment analysis (using GoNet).

p FDR: False discovery rate p correction.

**ESF, Table 12. Biological GO term classifications of a subset of downregulated differently expressed proteins in major depression**.

| **ID term** | **Term description** | **p** | **p FDR** | **Number of DEPs** | **Genes** |
| --- | --- | --- | --- | --- | --- |
| GO:1901214 | regulation of neuron death | 2.78E-07 | 1.88E-03 | 4 | CCL3\|CSF1\|IL10\|NGF |
| GO:0022603 | regulation of anatomical structure morphogenesis | 4.20E-07 | 1.88E-03 | 5 | CCL3\|CSF1\|IL10\|NGF\|VEGFA |
| GO:1904141 | positive regulation of microglial cell migration | 5.11E-07 | 1.88E-03 | 2 | CCL3\|CSF1 |
| GO:0002690 | positive regulation of leukocyte chemotaxis | 8.17E-07 | 2.25E-03 | 3 | CCL3\|CSF1\|VEGFA |
| GO:1904139 | regulation of microglial cell migration | 1.07E-06 | 2.25E-03 | 2 | CCL3\|CSF1 |
| GO:0051094 | positive regulation of developmental process | 1.41E-06 | 2.25E-03 | 5 | CCL3\|CSF1\|IL10\|NGF\|VEGFA |
| GO:0050769 | positive regulation of neurogenesis | 1.44E-06 | 2.25E-03 | 4 | CCL3\|CSF1\|NGF\|VEGFA |
| GO:0002688 | regulation of leukocyte chemotaxis | 1.75E-06 | 2.25E-03 | 3 | CCL3\|CSF1\|VEGFA |
| GO:1903977 | positive regulation of glial cell migration | 1.84E-06 | 2.25E-03 | 2 | CCL3\|CSF1 |
| GO:0051962 | positive regulation of nervous system development | 2.48E-06 | 2.41E-03 | 4 | CCL3\|CSF1\|NGF\|VEGFA |
| GO:0010720 | positive regulation of cell development | 2.55E-06 | 2.41E-03 | 4 | CCL3\|CSF1\|NGF\|VEGFA |
| GO:0002687 | positive regulation of leukocyte migration | 2.62E-06 | 2.41E-03 | 3 | CCL3\|CSF1\|VEGFA |

The differentially expressed proteins (DEPs) included are: CCL3, CSF1, VEGFA, NGF and IL10.

Results of Gene Ontology (GO) biological process functional enrichment analysis (using GoNet).

p FDR: False discovery rate p correction.

**ESF, Table 13. Biological GO term classifications of downregulated differently expressed proteins in major depression.**

| **ID term** | **Term description** | **p** | **p FDR** | **Number of DEPs** | **Genes** |
| --- | --- | --- | --- | --- | --- |
| GO:1904894 | positive regulation of receptor signaling pathway via STAT | 1.00E-10 | 4.48E-07 | 5 | IFNA2\|IL10\|IL12B\|IL13\|VEGFA |
| GO:0019221 | cytokine-mediated signaling pathway | 4.00E-10 | 1.32E-06 | 7 | IFNA2\|IL10\|IL12B\|IL13\|IL1A\|IL1B\|VEGFA |
| GO:0042981 | regulation of apoptotic process | 1.24E-07 | 7.59E-05 | 7 | IL10\|IL12B\|IL13\|IL1A\|IL1B\|NGF\|VEGFA |
| GO:0043067 | regulation of programmed cell death | 1.36E-07 | 7.80E-05 | 7 | IL10\|IL12B\|IL13\|IL1A\|IL1B\|NGF\|VEGFA |
| GO:0043066 | negative regulation of apoptotic process | 4.970E-08 | 9.846E-05 | 6 | IL10\|IL13\|IL1A\|IL1B\|NGF\|VEGFA |
| GO:0043069 | negative regulation of programmed cell death | 5.510E-08 | 9.846E-05 | 6 | IL10\|IL13\|IL1A\|IL1B\|NGF\|VEGFA |
| GO:0002700 | regulation of production of molecular mediator of immune response | 8.280E-08 | 9.846E-05 | 4 | IFNA2\|IL10\|IL13\|IL1B |
| GO:0060548 | negative regulation of cell death | 9.610E-08 | 9.846E-05 | 6 | IL10\|IL13\|IL1A\|IL1B\|NGF\|VEGFA |
| GO:0010628 | positive regulation of gene expression | 9.640E-08 | 9.846E-05 | 7 | IFNA2\|IL10\|IL13\|IL1A\|IL1B\|NGF\|VEGFA |
| GO:0071345 | cellular response to cytokine stimulus | 1.027E-07 | 9.846E-05 | 6 | IFNA2\|IL10\|IL13\|IL1A\|IL1B\|VEGFA |
| GO:2000026 | regulation of multicellular organismal development | 1.072E-07 | 9.846E-05 | 7 | IFNA2\|IL10\|IL13\|IL1A\|IL1B\|NGF\|VEGFA |
| GO:1901215 | negative regulation of neuron death | 4.30E-06 | 5.92E-03 | 3 | IL10\|IL13\|NGF |

See ESF, Table 4 for the downregulated differentially expressed proteins (DEPs) of principal component PCinv (IFNA2, IL1A, IL1B, IL12B, NGF, IL10, IL13, and VEGFA.

Results of Gene Ontology (GO) biological process functional enrichment analysis (using GoNet).

p FDR: False discovery rate p correction.
